# Accounting for assay performance when estimating the temporal dynamics in SARS-CoV-2 seroprevalence in the U.S

**DOI:** 10.1101/2022.09.13.22279702

**Authors:** Bernardo García-Carreras, Matt D. T. Hitchings, Michael A. Johansson, Matthew Biggerstaff, Rachel B. Slayton, Jessica M. Healy, Justin Lessler, Talia Quandelacy, Henrik Salje, Angkana T. Huang, Derek A. T. Cummings

## Abstract

Estimating the incidence of SARS-CoV-2 infection is central to understanding the state of the pandemic. Seroprevalence studies are often used to assess cumulative infections as they can identify asymptomatic infection. Since July 2020, commercial laboratories have conducted nationwide serosurveys for the U.S. CDC. They employed three assays, with different sensitivities and specificities, potentially introducing biases in seroprevalence estimates. Using mechanistic models, we show that accounting for assays explains some of the observed state-to-state variation in seroprevalence, and when integrating case and death surveillance data, we show that when using the Abbott assay, estimates of proportions infected can differ substantially from seroprevalence estimates. We also found that states with higher proportions infected (before or after vaccination) had lower vaccination coverages, a pattern corroborated using a separate dataset. Finally, to understand vaccination rates relative to the increase in cases, we estimated the proportions of the population that received a vaccine prior to infection.

## Introduction

Estimating the cumulative proportion of the population infected with SARS-CoV-2 is central to understanding the current state of the pandemic, assessing the susceptibility of the population, and to planning and targeting public health responses. Epidemiological models and other statistical approaches can be used to estimate cumulative infections using reported positive SARS-CoV-2 PCR tests, COVID-19 deaths, and other surveillance data (***Wu et al., 2020***; ***Chiu and Ndeffo-Mbah, 2021***; ***Irons and Raftery, 2021***; ***Lu et al., 2021***; ***Noh and Danuser, 2021***; ***Pei et al., 2021***; ***Sánchez-Romero et al., 2021***). Such studies revealed large underreporting of cases detected through case surveillance due to asymptomatic infections and limited laboratory testing. Seroprevalence studies based on a random sample of the population may be the gold standard for assessing the proportion infected but are expensive and logistically complicated to perform.

Since July 2020, commercial laboratories have conducted regular nationwide serosurveys for the CDC (***Bajema et al., 2021***; ***Clarke et al., 2022***). These surveys and other convenience and representative seroprevalence studies (***Havers et al., 2020***; ***Naranbhai et al., 2020***; ***Anand et al., 2020***; ***Menachemi et al., 2020***; ***Venugopal et al., 2021***; ***Bajema et al., 2021***; ***Lamba et al., 2021***; ***Bruckner et al., 2021***; ***Kline et al., 2021***; ***Kalish et al., 2021***; ***Jones et al., 2021***; ***Sullivan et al., 2022***; ***Routledge et al., 2022***; also see https://covid19serohub.nih.gov) have provided estimates of the cumulative proportion of the population with a history of at least one infection with SARS-CoV-2 in the United States at the national and local level. Modeling approaches have also used seroprevalence studies to improve estimates of critical parameters (e.g., the infection fatality rate) or to compare to model outputs (***Irons and Raftery, 2021***; ***Lu et al., 2021***; ***Chitwood et al., 2022***).

However, serosurveys can produce biased estimates of the proportion infected based on the samples and methods used. Convenience samples using existing serum and hospital-based samples may not be representative of the general population. Seroprevalence studies focusing on individuals seeking care for reasons unrelated to COVID-19, such as those conducted by the CDC, can underestimate the extent of mild infections due to tests being evaluated and calibrated mostly on patients with symptoms (***Burgess et al., 2020***; ***Takahashi et al., 2020***). Moreover, waning of antibodies to undetectable levels following infection has been observed (***Patel et al., 2020***; ***Ibarrondo et al., 2020***). Observed waning varies substantially between assays due to differences in their formats (e.g., whether the assays use direct or indirect detection formats; ***Macdonald et al., 2022***) and resulting variation in their sensitivities and specificities (***Peluso et al., 2021***; ***Bond et al., 2021***; ***Montesinos et al., 2021***; ***Takahashi et al., 2021***; ***Stone et al., 2022***). For example, when using manufacturer-recommended cutoff points to determine seropositivity, ***Peluso et al***. (***2021***) and ***Stone et al***. (***2022***) found lower sensitivities using ARCHITECT SARS-CoV-2 IgG immunoassay targeting the nucleocapsid protein (“Abbott”) than with Ortho-Clinical Diagnostics VITROS SARS-CoV-2 Total Ig and IgG (the latter only in Peluso *et al*.) immunoassay targeting the spike protein (“Ortho”) or Roche Elecsys Anti-SARS-CoV-2 pan-immunoglobulin immunoassay that targets the nucleocapsid protein (“Roche”). However, sensitivities to recent infections in ***Peluso et al***. (***2021***) were similar across all three assays. Both studies also estimated systematically faster waning using the Abbott assay while they found no evidence of waning for the Roche assay. As a result, all else remaining equal, antibody waning means that seroprevalence estimates will constitute an underestimate of the proportion infected.

In this study, we use CDC’s commercial laboratory nationwide serosurvey data and multiple other data sources to explain the observed spatio-temporal patterns in seroprevalence in the United States, with a particular focus on the role played by the different assays used, waning of antibodies, and the implications for estimating the proportion infected. We explore the impact of waning antibodies using a simple mechanistic model, where we adjust seroprevalence to estimate the proportion infected across the United States. Finally, to gain insight into the composition of sources of immunity, we compare the spatial patterns in estimated proportion infected with vaccination coverage across states over time.

## Results

We used data from CDC’s nationwide antibody serosurveys from commercial laboratories, which measures infection-induced seroprevalence. This study included both anti-nucleocapsid (anti-N) and anti-spike (anti-S) antibody assays prior to widespread vaccination campaigns, after which it included only anti-N assays. Anti-N assay seropositivity is reflective of prior infection with SARS-CoV-2 and not of vaccination with vaccines available in the United States, which contain only the spike protein; seroprevalence is also not a quantitative measure of current immunity status. We will henceforth refer to infection-induced seroprevalence as “seroprevalence”. Serosurveys started in July 2020 (round 1), and as of January 2022 (round 29), seroprevalence ranged from 18% in Vermont to 56% in Wisconsin. By then, the proportions of state populations reported as confirmed COVID-19 cases ranged from 10% in Hawaii to 25% in Rhode Island, and the proportions for confirmed deaths ranged from 0.1% in Vermont to 0.4% in Mississippi, with marked heterogeneity across states by round (e.g., Figure S1 in supporting information (SI)). Rank order of states by seroprevalence at a point in time differed quite markedly from that by proportion of the population reported as a case (***Figure 1***). To explain spatio-temporal variation in seroprevalence across states, we fit two sets of models. The first model (“reference model”) includes cumulative proportions of populations reported as cases and deaths as explanatory variables while accounting for a range of other factors including the assays used in each survey. The second set of models (“waning models”) explicitly incorporates the temporal effect of different waning rates (depending on the assay being used) on seroprevalence estimates (see Methods and Materials). Comparison of the waning models with the reference model enables assessment of the proposed mechanistic model of waning in measured antibodies and its relative ability to explain observed patterns.

**Figure 1.**
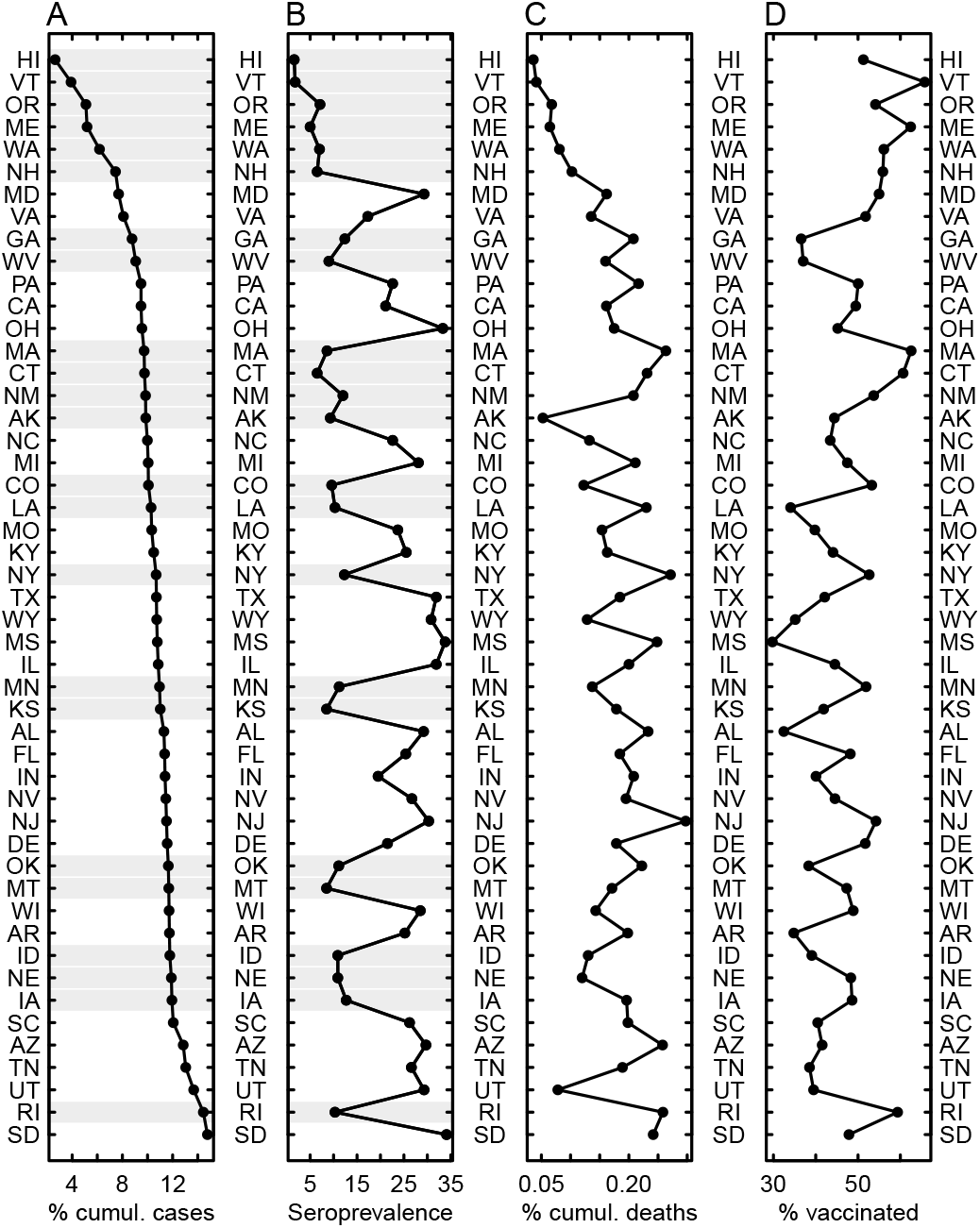
States ranked by different metrics. Ranked (A) cumulative percentage of the population reported as a COVID-19 case, (B) seroprevalence from the CDC nationwide serosurveys, (C) cumulative percentage of the population reported as a COVID-19 death, and (D) vaccination (with a full series) coverage for July 2021 (round 24, the last round before the Roche assay started being used exclusively). Gray shading in A and B show serosurveys that at that point in time exclusively used the Abbott assay.

### Variation in infection-induced seroprevalence associated with the use of different assays

Seroprevalence varied systematically as a function of the specific assays used in the surveys (Abbott, Ortho, or Roche). In the reference model, higher proportions of the Abbott assay were associated with lower seroprevalence while Roche assay use was associated with higher seroprevalence (Table S1 and Figure S2 in SI; the Ortho assay was included in the model as the comparison group).

As a result, some of the spatial variation observed in the nationwide serosurveys was attributable to the spatially heterogeneous use of assays (Figures S3–S6 in SI). Using the reference model, we estimated the seroprevalence that would have resulted had all states exclusively used one of the three assays alone. We estimated that if the serosurveys had exclusively used the Roche assay (highest seroprevalence), estimated seroprevalence country-wide could have been 21 percentage points higher in January 2022 than if only the Abbott assay (lowest seroprevalence) was to have been used (Figure S7 in SI). There was substantial state-to-state variation in the change in expected seroprevalence had the Roche assay been exclusive used; for example, seroprevalence would have been over 27 percentage points higher in Iowa in May 2021 (round 21) had they exclusively used the Roche assay, relative to the actual survey estimates (Figure S7 in SI).

### Accounting for waning helps explain variation in infection-induced seroprevalence

Accounting for waning detection of antibodies over time improved model fit over the reference model across various metrics (***Figure 2*** and Figure S8 in SI; ***Table 1***). Results support a faster waning rate for the Abbott assay, while there was no clear evidence of waning in the Roche assay. Time to seroreversion (the time for antibodies to fall below a detection threshold) in the Ortho assay was found to be longer than that for the Abbott assay, but because the Ortho assay was only used up to January 2021 (except in Puerto Rico, not included in this study), we could only explore up to a maximum of 49 weeks of time to seroreversion. The best fitting models by root-mean-square error (RMSE) and median leave-one-out (LOO) RMSE had RMSEs and median LOO RMSEs approximately 0.91 and 0.90 times that of the reference model without waning, respectively. The best model by LOO RMSE had a mean time of seroreversion of 24 weeks for the Abbott assay, 42 weeks for the Ortho assay, and there was no evidence of seroreversion in the Roche assay over the time period studied (97 weeks), with cases seroconverting one week prior to being reported as a case (-1 week detection delay; ***Table 1***). Note, however, that there was uncertainty around these parameter estimates (***Figure 2***). The best models by Akaike information criterion (AIC) and RMSE were similar, with estimated Abbott assay time to seroreversion of 39 and 31 weeks, respectively, and no clear evidence of waning in the Ortho assay (with a time to seroreversion ≥ 49 weeks; ***Table 1***). After accounting for waning, the negative and positive associations between proportion of Abbott and Roche assays with seroprevalence remained, albeit with smaller effect sizes (Table S1), potentially implying lower sensitivity of the Abbott assay for detecting recent infections relative to the other two.

Next, we used the best waning model to estimate the proportion infected by correcting the seroprevalence for assay use and seroreversion (see Methods and Materials) and compared it to reported seroprevalence. The difference between estimated proportions infected and seroprevalence is greatest in states and time points in which the Abbott assay was predominantly used (e.g., ***Figure 3*** and Figures S4–S6 in SI). The estimated proportion infected was at least 10 percentage points higher than the seroprevalence in six states in January 2021, and in 17 states in July 2021, although when averaged across the country (using state populations as weights), the difference was at most 5 percentage points (***Figure 3***). Our results also show that the decreasing seroprevalence over time observed in some states was at least in part attributable to the assays used and corresponding waning rates (e.g., most of the states shown in ***Figure 3***). The resultant time series of proportion infected differed not only quantitatively but in some cases also qualitatively from the seroprevalence estimates (***Figure 3*** and Figure S9 in SI).

**Table 1.** Best waning models across three metrics, compared to the reference model. Metrics for the best models by Akaike information criterion (AIC), root-mean-square error (RMSE), and leave-one-out (LOO) median RMSE, compared to metrics for the reference model (with no waning). For example, the column “AIC” indicates the best waning model chosen by AIC (see ***Figure 2***). AIC values for waning models account for the added parameters being selected (times to seroreversion and detection lead or lag). ΔAIC values in the table are relative to the lowest AIC in the models shown (the best fitting waning model by AIC).

|  | Best fitting waning model, by each metric |  |  | Reference model |
| --- | --- | --- | --- | --- |
|  | AIC | RMSE | LOO median RMSE |  |
| Detection lead or lag (weeks) | 0 | 0 | −1 | − |
| Abbott time to seroreversion (weeks) | 39 | 31 | 24 | − |
| Ortho time to seroreversion (weeks) | 49* | 49* | 42 | − |
| Roche time to seroreversion (weeks) | 97* | 97* | 97* | − |
| ΔAIC | 0 | 54 | 215 | 679 |
| RMSE | 0.0203 | 0.0198 | 0.0199 | 0.0217 |
| LOO median RMSE | 0.0207 | 0.0210 | 0.0204 | 0.0223 |
| Observations | 1398 | 1398 | 1398 | 1398 |
| Residual degrees of freedom | 1285 | 1285 | 1285 | 1285 |

\* These are lower bounds because they are the maximum number of weeks for which these assays could be evaluated. The Ortho assay was only used, to a limited extent, in the first 13 rounds of the nationwide serosurveys (until January 2021), while the Abbott assay was used in the first 24 rounds (until July 2021; see Methods and Materials and Figures S1 and S3 in SI).

**Figure 2.**
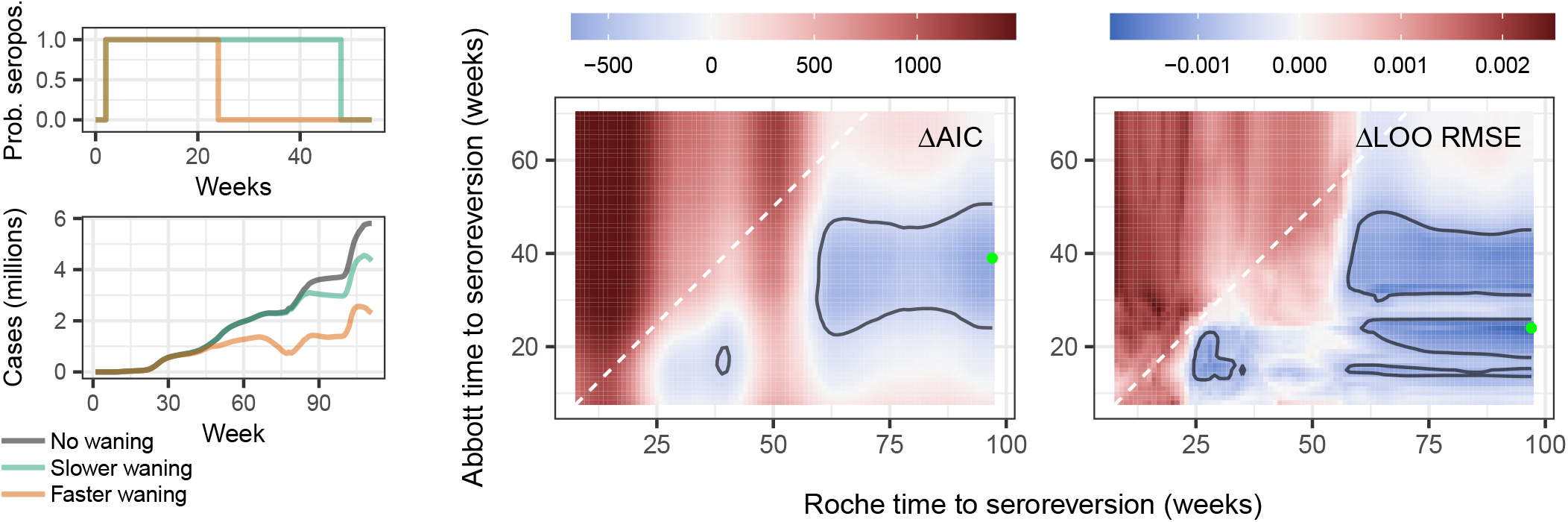
Rationale behind the mechanistic models, and comparison of waning models with the reference model across two metrics. Inclusion of waning helps explain patterns in seroprevalence better. Left panels explain how we mechanistically incorporate waning into the models. A case is expected to test positive in a survey for a limited amount of time before seroreverting (top left), leading to different waning patterns that depend on the time to seroreversion (bottom left; see Figure S14 in SI). In our models we use three different times to seroreversion, one for each assay. Within each tile panel, each pixel corresponds to a single model, where cases have been adjusted assuming three different times to seroreversion, one for each assay. The best models had a case seroconvert in the same week (by AIC) or one week before being reported (by leave-one-out median RMSE; Table 1). Tile plots assume the best model’s time to seroreversion for the Ortho assay (49 weeks by AIC, 42 weeks by LOO median RMSE), and show model performance for the remaining two variables, the times to seroreversion for Roche (x-axes) and Abbott (y-axes) assays. The two tile plots show results for two different model metrics: AIC, and LOO median RMSE. Metrics are expressed relative to the metric for the reference model (that does not account for waning); blues (respectively reds) indicate waning models that are better (respectively worse), per that metric, relative to the model without waning. Contour lines in the tile plots enclose the best five percentile models for each metric. Green points indicate the best model by each metric, and contour lines enclose the best five percentile models as per each metric. See Table 1 for the corresponding best waning model by each metric, and Figure S8 in SI for more complete results.

**Figure 3.**
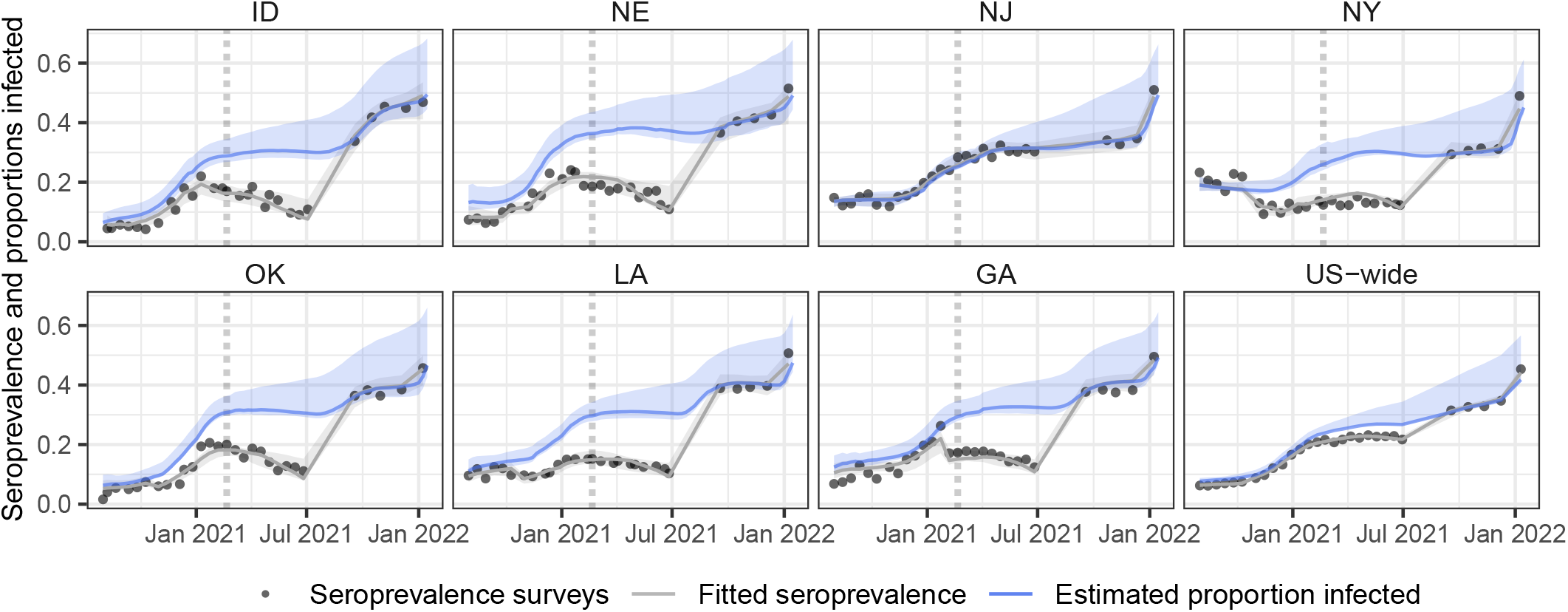
Time series of survey seroprevalence and estimated proportions infected for seven example states and U.S.-wide. Example time series of survey seroprevalence, fitted seroprevalence and estimated proportion infected, for states for which seroprevalence was estimated primarily using the Abbott assay prior to September 2021 (see Figures S1 and S3 in SI), except for New Jersey, for which the Roche assay was exclusively used, and U.S.-wide estimates (bottom right). The proportion infected was estimated using the best waning generalized linear model (GLM) by LOO median RMSE (***Figure 2***; ***Table 1***). U.S.-wide estimates were obtained by taking mean values per round weighted by population, and each round was plotted taking the mean week for that round across all states. Uncertainty envelopes around fits and estimated proportions infected include model uncertainty and uncertainty around the selection of times to seroreversion and lead or lag between seroprevalence and reported cases. The U.S.-wide ribbon does not include model uncertainty. Vertical dotted lines indicate the start of the vaccination campaigns. See Figure S9 in SI for time series for all states included in the model.

### Spatial heterogeneity in infection-induced seroprevalence, estimated proportion infected, and vaccination coverage

Some of the observed spatial heterogeneity in seroprevalence (***Figure 4A***) was a result of the use of different assays and their associated waning rates (Figures S3–S6 in SI). For example, by July 2021 (round 24, the last round prior to the Roche assay being used exclusively), the standard deviation in percentage seroprevalence across states was 10% (***Figure 4A***), while after correcting for assay use and waning (***Figure 4B***), it was 7.2% (also see Figure S10 in SI). We also found that as vaccine distribution increased in 2021, states with higher vaccine coverage were associated with lower estimates of the proportion infected (***Figure 4E***). We combined the estimates of the proportion infected from our best waning model with the vaccination coverage (thus including seropositives Seroprevalence and proportions infected from both natural infection and/or vaccination, which we henceforth refer to as the estimated proportion infected and/or vaccinated, or EPIV), by assuming that the probabilities of being infected and vaccinated with a complete series are independent. The differences between states were further reduced when considering EPIV (***Figure 4***, comparing maps B and D; Figure S10 in SI); in July 2021, the standard deviation in the EPIV was 5.6% (Figure S10 in SI). A comparison of time series in individual states (***Figure 4F***) illustrates the relationship between vaccine coverage and the estimated proportion infected over time. Of note, Washington, a state with low estimated proportion infected pre-vaccination and higher vaccination coverage maintained a low proportion infected post-vaccination, while Alabama had high estimated proportion infected pre-vaccination and achieved lower vaccine coverage.

**Figure 4.**
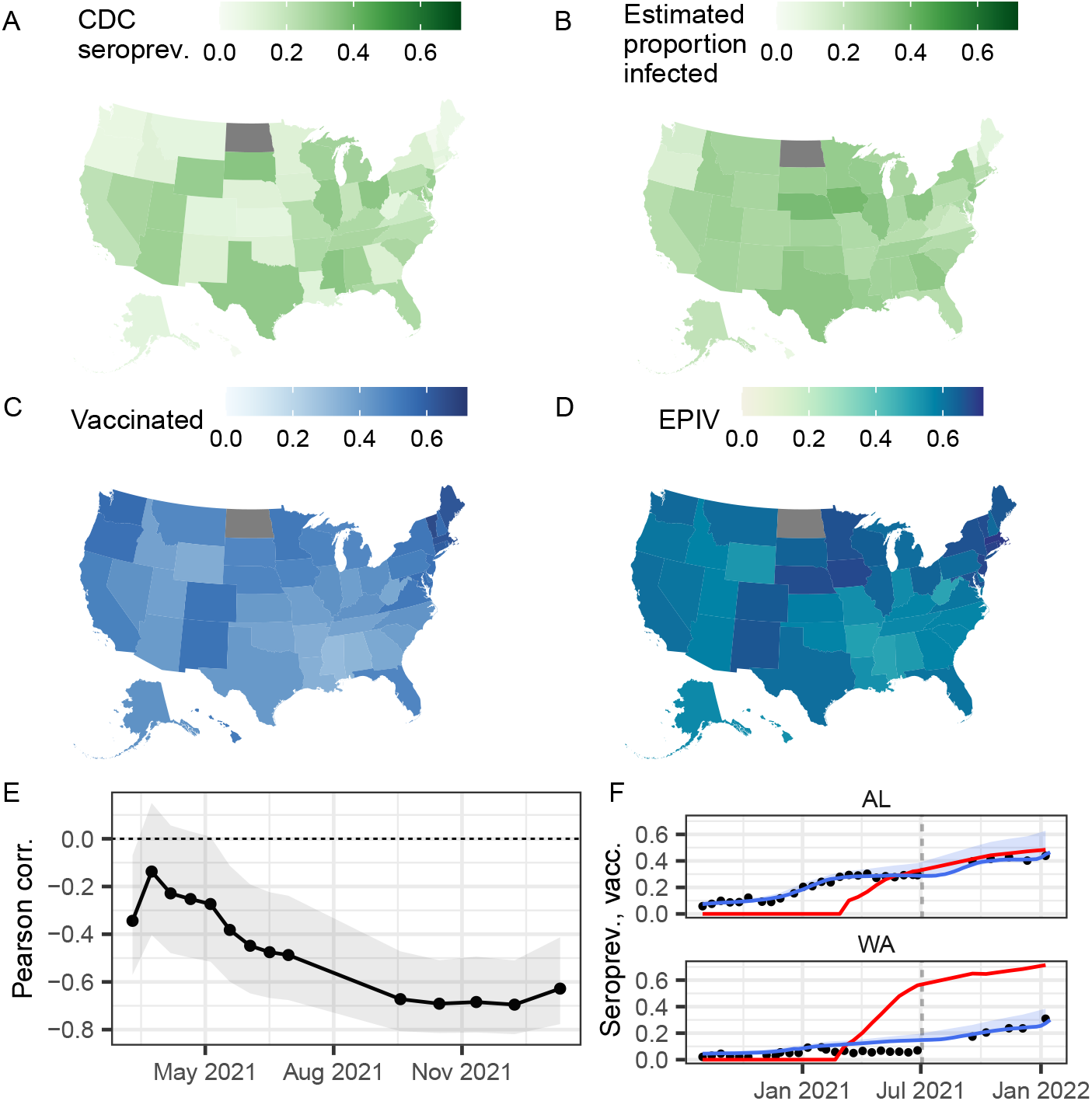
Spatial patterns and correlations. Spatial variation in July 2021 (round 24) in (A) CDC nationwide serosurvey seroprevalence, (B) estimated proportion infected, (C) proportion of the population with a complete series of a vaccine, (D) estimated proportion infected and/or vaccinated (EPIV; assuming independent probabilities of having had a natural infection and being vaccinated), (E) round-by-round Pearson correlation between the proportion of the population vaccinated and the estimated proportion infected (shaded areas show the 95% uncertainty intervals for a two-sided test), and (F) example time series of seroprevalence estimates (black points), estimated proportion infected (blue lines; shaded areas show the uncertainty intervals), and proportion of the population vaccinated with a full series (red lines) for two states. Panels A–D show maps for July 2021 (round 24); its point in time is shown in panel F as vertical gray dashed lines. In panel E, a negative correlation means that states with a higher vaccination coverage tended to be those with lower proportion infected. See Figure S9 in SI for time series like those in panel F for all states.

### Pre-infection vaccine coverage

The greatest public health benefit of vaccines is likely achieved when administered to individuals prior to infection. Maximizing the vaccine coverage of individuals pre-infection would have required both limiting, to the extent possible, transmission (e.g., by implementing non-pharmaceutical interventions), and an effective vaccination campaign. Our reconstructions of the proportion infected show that the degree to which transmission was constrained in the United States varied across states and over time (***Figure 3*** and Figure S9 in SI), by more accurately showing when the cumulative proportion infected remained flat. Furthermore, although vaccination campaigns started almost simultaneously across the country, differences across states in vaccination rates and coverage quickly emerged (Figures S5 and S6 in SI). To give insight on the coverage and speed of the vaccination campaign relative to the speed at which cases increased, we estimated the proportion of the total population that was vaccinated with a complete series of doses before being infected, assuming that vaccinations were distributed independently of prior infection status. The proportion of the whole population who were vaccinated and not previously infected ranged from 6% in Utah to 15% in Alaska in mid-March 2021, before widespread availability of vaccination to individuals over ages 65 years, and from 21% in Idaho to 42% in Vermont by mid-January 2022 (Figure S11 in SI).

### Comparison with an independent dataset

Finally, we compared estimates produced by our models with an independent dataset, the nationwide blood donor serosurvey (***Jones et al., 2021***). Infection-induced seroprevalence estimates from the two sets of surveys are clearly correlated (Pearson correlation of 0.85), albeit with substantial variation (Figure S12 in SI), while our estimated proportion infected was also highly correlated to the blood donor serosurvey estimates (Pearson correlation of 0.93), although our estimates tended to be higher.

The blood donor surveys included both anti-N and anti-S assays, allowing estimates of seropositivity from both natural infection and vaccination, respectively (***Jones et al., 2021***). Our EPIV values were substantially lower than the anti-S seroprevalence estimated in the blood donor survey (***Figure 5***), especially after vaccinations started. Including individuals with at least one vaccine dose in the EPIV brought our values closer to the blood donor survey estimates. When assuming a perfect negative correlation between having one or more doses of a vaccine and having ever tested positive due to an infection (adding the two proportions, thus constituting an upper limit), our EPIV were comparable to those of the blood donor surveys. It is, however, also important to note that differences between our EPIV estimates and the blood donor surveys could also be attributable in part to the uncertainty around the time to seroreversion of, particularly, the Roche assay (***Figure 2*** and 5).

**Figure 5.**
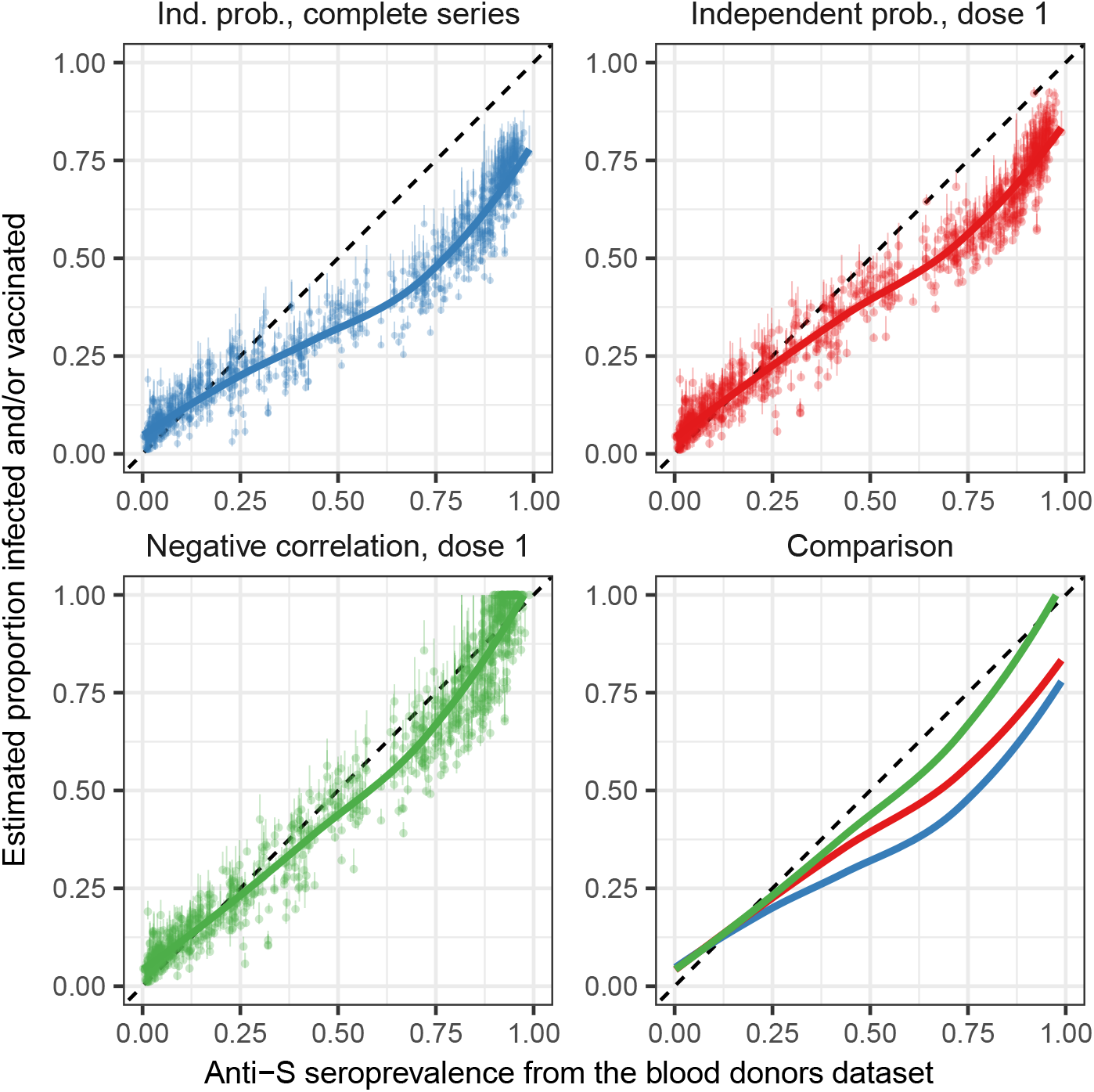
Comparison of our estimates of proportions infected and/or vaccinated with the blood donors anti-S seroprevalence estimates. Comparing estimated proportions infected and/or vaccinated (EPIV) with estimated anti-S seroprevalence from blood donor samples. The seroprevalence from the blood donor samples includes both infected and vaccinated individuals, and the EPIV combines estimated proportion infected (adjusting for assay use and waning) with the proportion vaccinated. Lines are LOESS fits to the data points shown, and vertical lines show uncertainty in EPIV values due to that around our estimated proportions infected. The blue line and points assume that the probability of infection and being vaccinated (with two doses) are independent, red lines and points use the proportion vaccinated with at least one dose, while the green line and points use the proportion vaccinated with at least one dose, and assume a perfect negative correlation between vaccination coverage and estimated proportion infected. See Figure S13 in SI for comparisons of time series of these quantities per state.

## Discussion

Our results show that heterogeneous spatio-temporal patterns in seroprevalence are in part explained by which assays were used in the surveys: seroprevalence was lower in states that made greater use of the Abbott assay, underlining variation in the ability to detect infection across assays, and possibly indicative of the different detection thresholds that define seropositivity used by each assay. Additionally, the fact that models incorporating waning better fit the data than the reference model suggests that the temporal aspects of waning in antibodies are important in explaining patterns in seroprevalence; waning rates in seroprevalence studies were assay dependent. The times to seroreversion were found to be likely distinctly lower in the Abbott assay than in the Ortho or Roche assays (for the latter in particular, no strong evidence of waning was found at a population level); these differences are supported by other literature (e.g., ***Peluso et al., 2021***) and likely related to characteristics of the assay target. Using our estimates of times to seroreversion for the different assays, we show that estimated proportions infected differ quantitatively and qualitatively from seroprevalence in states that made substantial use of the Abbott assay. Accounting for the assays used reduces differences across states in seroprevalence and suggests a more homogeneous impact of the pandemic across the United States than would otherwise be surmised based on the surveys alone. This result is confirmed by the lower spatial variation in seroprevalence from September 2021 onwards, when all states used the same (Roche) assay. The remaining variation across states was further reduced when also accounting for vaccination rates; the estimated proportions infected were negatively correlated with vaccination rates across states.

To highlight the influence of choice of assays on seroprevalence estimates, we compare the seroprevalence estimated in New York and New Jersey. Both states experienced qualitatively similar outbreak dynamics according to reported cases and deaths, yet the surveys produced very different seroprevalence estimates (both in their absolute values, and in particular their evolution over time; see Figure 3). The maximum difference in their seroprevalence was 19 percentage points (13% in New York, 32% in New Jersey in May 2021). Seroprevalence in New York exhibited a conspicuous drop between October and November, a drop that was not observed in neighboring New Jersey. Our results show that the drop in New York can at least in part be explained by a switch from using the Roche assay to a mix of the Ortho and Abbott assays in October 2020 to exclusively using the Abbott assay by January 2021 to produce the New York seroprevalence estimates. However, sampling for the study was also changed in November 2020 to include a larger proportion of specimens from outside the New York City metropolitan area, which had experienced the largest spike in early cases. Estimates for New Jersey were obtained exclusively using the Roche assay. Accounting for this difference produces estimates of proportion infected that are more similar in magnitude and in trend across the two states. For instance, the maximum difference between the two states after adjusting for assay use is less than 6 percentage points, in line with the maximum difference reported in seroprevalence after July 2021 (when both used the Roche assay) of just over 4 percentage points.

Longitudinal studies quantifying within-individual antibody kinetics have also previously shown (albeit with relatively small sample sizes) how antibody levels and waning rates can markedly vary depending on the assay used and on disease severity (***Peluso et al., 2021***; ***Stone et al., 2022***, but also see ***Arkhipova-Jenkins et al., 2021***). ***Peluso et al***. (***2021***) found similar mean times to seroreversion for the Abbott assay (23 and 33 weeks for non-hospitalized and hospitalized individuals, respectively), compared to 24 weeks in our best waning model, and 39 and 79 weeks for the Ortho assay compared to 42 weeks in our results (although note that the Ortho assay was phased out by January 2021), while they found no evidence of waning for the Roche assay, and neither did we up to 97 weeks, albeit with significant uncertainty around our estimate. ***Stone et al***. (***2022***) also reported distinctly faster waning rates in the Abbott assay. Moreover, even after accounting for differential waning between assays, Abbott assay use was associated with lower seroprevalence, possibly suggesting lower sensitivity to recent infections, although this finding was not consistent with ***Peluso et al***. (***2021***), who found similar sensitivities to recent infections across all three assays.

Previous studies have also leveraged individual-level immune dynamics to produce corrected seroprevalence estimates. For instance, using time series of reported cases, deaths, or hospitalizations, ***Takahashi et al***. (***2021***) used time-varying assay sensitivities (and their variation with disease severity, estimated in individual-level data) to produce adjusted estimates of seroprevalence across five locations. Where they incorporated individual-level data into their methodology, we recovered individual-level patterns across a large population.

The spatial heterogeneity observed in the nationwide serosurveys was to an extent attributable to assays used and waning; variation across states in the estimated proportion infected was distinctly lower. Nevertheless, the vaccination campaign started at a point in time when the estimated proportion infected still differed by >34 percentage points across states, and this maximum range grew to >39 percentage points by January 2022. This means that vaccination campaigns started on a relatively heterogeneous landscape of immunity and that heterogeneity increased with vaccinations and subsequent infections. Uptake of vaccines also varied across states, with the proportions of the population vaccinated differing across states by as much as 30 percentage points by January 2022. Vaccination coverage was negatively correlated with the estimated proportion infected, a finding corroborated in a comparison with an independent dataset. This negative correlation implies that differences among states in the proportions of state populations that have experienced an immune response (whether by infection or vaccination) is lower than expected based on vaccination coverage or the proportion infected alone. However, the negative correlation also suggests that the composition of the source of immunity (from either infection or vaccination) is likely heterogeneous across states. Consequently, were immune protection from infections and vaccines found to differ systematically (and there are indications that this may indeed be the case, e.g., ***Goldberg et al., 2022***; ***Carazo et al., 2022***), the result of future waves of the pandemic may also be expected to be spatially heterogeneous. We also presented a metric meant to capture the rates at which states delivered vaccines in relation to the rate at which cases accrued. Reconstruction of the dynamics of cumulative infections allows for greater investigation of heterogeneity between locales that might be used to guide future public health responses.

The differences in our estimates of the proportion infected and estimated proportion infected and/or vaccinated (estimates without and with vaccination, while accounting for assay use and waning) with the blood donors surveys could in part be attributed to likely differences in the biases in the sampling inherent to the two surveys. The nationwide serosurveys that form the basis of our estimates use samples from individuals seeking medical care for reasons unrelated to COVID-19. On the other hand, people who donate blood may differ from the overall population in important ways; for instance, blood donors are more likely to be healthy, non-pregnant adults, certain groups (e.g., younger age categories) may be systematically underrepresented, and for example, their vaccination uptake might be systematically higher (e.g., ***Busch et al., 2022***). The assays used were also not the same across the two sets of serosurveys. Nonetheless, the comparison supports the negative correlation between proportion infected and vaccination coverage.

A number of caveats should be taken into consideration when interpreting our results. As noted above, our results are based on serosurveys using a convenience sample of individuals that sought health care for reasons other than COVID-19; this sample could deviate from the wider population in important ways and not be representative. For example, this group may have experienced different rates of severe illness upon infection with SARS-CoV-2, an important determinant of immune response, than the general population (***Takahashi et al., 2020, 2021***; ***Peluso et al., 2021***) and may have systematically different healthcare seeking behavior. Furthermore, biases in the sampling could also vary over time and across states, for instance as a function of the numbers of cases and underlying demographics. Rates of seroconversion and reversion might also be different preand post-vaccination (e.g., ***Follmann et al., 2022***; ***Goldberg et al., 2022***; ***Carazo et al., 2022***). We use numbers of tests that were positive and negative in the models, meaning that we do not make explicit adjustments for race, ethnicity, age, or sex, although these factors are, to an extent, captured in the model with state-specific intercepts. It is also important to note that the serosurveys aim to determine evidence of prior infection by detecting the presence of IgG in samples, and as a result, our estimates of the proportion infected, too, focus on estimating prior infections. However, these do not necessarily reflect protection from new infection. Finally, to calculate the estimated proportion infected and/or vaccinated, we assumed that the probabilities of being infected and vaccinated were independent. However, vaccination may be associated with prior infection and the comparison to blood donor seroprevalence suggests that vaccination may be negatively correlated with the probability of prior infection.

The lags between seroconversion and a case being reported were, a priori, an important parameter to consider in accounting for potential systematic shifts in the time series of seroprevalence and reporting of cases and deaths. However, this parameter should be interpreted with caution. Serosurveys were conducted roughly every two to four weeks, and they reported time windows (that can be two to four weeks long) over which the surveys were performed; we here use the midpoints of the windows. Reporting delays might also be expected to vary over time, possibly as a function of the number of cases being reported. Nevertheless, our results do not provide strong evidence for a specific lead or lag, and this is reflected in the uncertainty estimates we provide.

Serosurveys will continue to be critical tools to understand determinants and predictors of infection, reinfection, duration of protection, antigen-specific protection to SARS-CoV-2 variants, and the underlying determinants of burden (e.g., the infection fatality ratio). Given the changing relationship between reported cases and infections due to reinfections and breakthrough cases and the increasing availability of at-home testing, statistical and mechanistic approaches to analyzing serosurvey data will become more important. Our results identifying signals of waning and the correlation between vaccination and prior infection suggest that large scale, aggregate datasets like the U.S. serosurveys may yield useful inferences on the relationships between serological responses, protection, and reinfection. However, further work will be needed to interpret serology as seropositivity saturates in the population and more individuals experience multiple immunizing events (i.e., re-infection, vaccine boosts).

## Methods and Materials

In this study we aimed to explain spatio-temporal variation in seroprevalence using binomial generalized linear models (GLMs). We included as covariates the variable use of different assays across time and space and assessed the evidence to support differential waning of seropositivity across assays. The correlations between the covariates used in the models are shown in Figure S14 in SI.

### Data

The serosurveys, conducted by the CDC and commercial laboratories, included samples obtained for reasons unrelated to COVID-19. Nationwide seroprevalence studies using available serum specimens (henceforth referred to as “nationwide serosurveys”) were conducted from July 2020, with the aim of estimating seroprevalence from infection per state approximately every two to four weeks (Bajema *et al*. 2021). The surveys are ongoing, but we here analyze surveys up to January 2022 (round 29). Three immunoassays were used: the Roche Elecsys Anti-SARS-CoV-2 pan-immunoglobulin immunoassay that targets the nucleocapsid protein (henceforth referred to as “Roche”), the Abbott ARCHITECT SARS-CoV-2 IgG immunoassay targeting the nucleocapsid protein (henceforth referred to as “Abbott”, and Ortho-Clinical Diagnostics VITROS SARS-CoV-2 IgG immunoassay targeting the spike protein (henceforth referred to as “Ortho”). Further details about the laboratory methods, including the sensitivity, specificity, can be found in Section “Laboratory methods” in SI. As all vaccines available in the United States generate antibodies to the spike protein only (anti-S), serosurveys conducted following the widespread availability of vaccines used exclusively assays measuring anti-N IgG. The Ortho assay measured anti-S antibodies, but their use was phased out in the states analyzed here by the end of January 2021, prior to the start of widespread vaccination campaigns. The nationwide serosurvey data were downloaded from the CDC (https://data.cdc.gov/Laboratory-Surveillance/Nationwide-Commercial-Laboratory-Seroprevalence-Su/d2tw-32xv). We assumed the surveys for each round took place on the middle date of the range given. We then used the number of positive and negative tests produced in each survey as our outcome variable. Because there were few completed survey rounds for North Dakota, it was excluded from analyses.

County-level daily laboratory-confirmed COVID-19 cases and deaths in the counties of the United States were downloaded from USAFacts (https://usafacts.org/visualizations/coronavirus-covid-19-spread-map/) on March 2, 2022. After aggregating the numbers of cases and deaths per state, and differencing the cumulative curves to obtain numbers of cases and deaths per day, we found negative values of both reported deaths and cases. If the negative value was immediately followed by the same (positive) value, those counts were canceled out. Otherwise, the negative total was discounted from previous days’ totals. We then aggregated numbers by week, recalculated cumulative numbers, and divided them by the respective state populations to produce cumulative percentages of the population that were reported as COVID-19 cases and deaths, for each nationwide serosurvey round. These data did not include Puerto Rico, so Puerto Rico is not included in our analyses.

Excess deaths data for each state were downloaded from the CDC (https://www.cdc.gov/nchs/nvss/vsrr/covid19/excess_deaths.htm). We kept the weighted data only (which attempts to correct for reporting delays). To estimate the number of (excess) deaths not attributable to COVID-19, we took the difference between excess deaths and reported deaths. We then calculated this number as a percentage of the state population.

Laboratory testing (PCR) time series per state were downloaded from HealthData.gov (https://healthdata.gov/dataset/COVID-19-Diagnostic-Laboratory-Testing-PCR-Testing/j8mb-icvb), and from these we estimated the cumulative number of tests performed relative to each state’s population up to each serosurvey round.

We used data on the distribution of assays used in each serosurvey round (***Wiegand et al., 2022***). The information provided included the number of tests, for each survey round, that were performed with each of three different assays (Abbott ARCHITECT IgG anti-N, Ortho VITROS IgG antiS, and Roche Elecsys Total Ig anti-N). From September 2021 (round 25) onwards, states switched to exclusively using the Roche Elecsys assay.

COVID-19 vaccination data were downloaded from the CDC (https://data.cdc.gov/Vaccinations/COVID-19-Vaccinations-in-the-United-States-Jurisdi/unsk-b7fc). We produced percentages of the populations that had been vaccinated with at least one dose of a vaccine, or with a complete series of the vaccine (individuals with a second dose of a two-dose vaccine or one dose of a single-dose vaccine) at each point in time per state. For a couple of states (e.g., Kentucky and West Virginia), vaccination coverage is not a monotonically increasing function of time. However, it is unclear from the data documentation what the reason for this pattern may be.

We downloaded data on COVID-19 hospitalizations from HealthData.gov (https://healthdata.gov/Hospital/COVID-19-Reported-Patient-Impact-and-Hospital-Capa/g62h-syeh). We calculated the cumulative total number of confirmed adult hospital admissions, and then obtained the percentage of the cumulative number of cases that had been hospitalized, per state.

The proportion of COVID-19 cases reported in different age categories was estimated using the CDC restricted access case surveillance line-list data (https://data.cdc.gov/Case-Surveillance/COVID-19-Case-Surveillance-Restricted-Access-Detai/mbd7-r32t). The reporting times in the CDC line list data are not expected to match those from USAFacts.gov. We assumed that the proportions of total cases being reported in each of the age categories was unlikely to undergo very rapid changes over time, so we estimated these proportions based on five-week rolling means of the cumulative number of total cases and cases reported in each age category.

We compared our estimates of proportions infected with a separate serosurvey conducted by the CDC: the nationwide blood donor serosurvey (https://covid.cdc.gov/covid-data-tracker/#nationwide-blood-d The survey estimates the proportion of the population with antibodies against SARS-CoV-2 (both anti-N and anti-S Ig), for which they used the Roche Elecsys Total Ig and Ortho VITROS Total Ig assays. Multiple estimates were provided for different parts of some states; we took the mean seroprevalence weighted by the number of tests to get a single estimate by state. Surveys were not necessarily performed in the same weeks as the nationwide serosurveys. To maximize the data used when comparing the two datasets, if surveys in the two datasets were performed one week before or after the other, the two values were still matched.

We square-root-transformed the cumulative percentages of the populations reported as cases and deaths, and the percentages of the populations hospitalized, because their distributions were heavily skewed and are expected to be the result of a multiplicative process. For similar reasons, we natural-log-transformed the percentage of state populations that were PCR tested. Models with these transformations performed better across model metrics used in this study than models without transformations.

The data used to fit the models is provided in SI Dataset S1.

### Models

We used binomial generalized linear models (GLMs). In all models, the number of positive and negative tests were the response variable. The “reference model” included an interaction between week and state (which aimed to capture changes in the percentages of COVID-19 infections reported as cases over time and across states); the square-root-transformed cumulative percentages of the state populations reported as a case and as a death, the percentage of the population reported as excess (unaccounted for) deaths, natural-log-transformed percentage of the population that had been tested (PCR), the cumulative percentage of the population that had been vaccinated, the square-root-transformed cumulative percentage of cases that had been hospitalized, the percentage of survey tests that utilized the Abbott and Roche assays (as two separate variables; we did not include a covariate for Ortho use because its inclusion would have been redundant, given percentages across the three assays always equal 100), and the percentage of cases being reported for different age categories (we did not include ages >70 years category as the inclusion would have been redundant given percentages across age categories add to 100). We weighted the model to account for the different proportions of the state populations that were tested in the nationwide serosurveys by scaling the positives and negatives such that the probability that an individual was tested was the same across all states (by using the state population divided by the mean state population as weights in the model).

While accounting for the seroprevalence associated with the use of different assays, the reference model above does not explicitly account for the waning in antibodies over time as quantified by each of the assays. As a point of comparison, we separately fit a suite of GLM models (henceforth, “waning models”) based on the reference model above, but which assumed a range of different antibody waning rates per assay. We proceed with the following strategy. The time series of the numbers of cases in a location is a (monotonically) increasing function of time, while the time series for seroprevalence need not be (if, for instance, antibodies did wane below detectability). We therefore “adjust” the reported cases to incorporate waning following a series of assumptions.

To adjust cases, we multiply each reported case by a step function produced using two parameters: (i) a lead or lag between a case seroconverting and it being reported; and (ii) a limited time during which that case would test positive before seroreverting (Figure S14 in SI). This produces an alternative “adjusted” time series of reported cases. Furthermore, we could hypothesize that different assays have potentially different times to seroreversion, and thus use multiple step functions to produce these adjusted time series of cases, one for each assay. We produce adjusted numbers of reported cases based on the proportions of each assay used to produce each seroprevalence estimate. We then fit different GLM models for all combinations of the three step functions (or waning rates). An improvement in model fit over the reference model provides evidence for the input waning rates. In this way, the impact assays have on seroprevalence is split into two components: the temporal waning rate, and the average seroprevalence associated with each assay, after accounting for waning, which can be interpreted as a proxy for assay sensitivity for recent infections. Note that while the Roche assay has been used through all survey rounds, the Abbott assay was used until July 2021 (round 24), and the Ortho assay was used until January 2021 (round 13). This means the maximum times to seroreversion we can explore for the Abbott and Ortho assays are 70 and 49 weeks, respectively, relative to the start of the pandemic.

We evaluated models using the Akaike Information Criterion (AIC), root-mean-squared-error (RMSE), and a leave-one-out (LOO) median RMSE. RMSE values were estimated by comparing model predictions on the response scale with nationwide serosurvey estimates. For the LOO RMSE, each round of the surveys was left out in turn, the model fit to the remaining rounds and used to predict the round left out. We then estimated the median RMSE from the predictions of the rounds left out. We estimate the proportion infected by taking the best waning model, and replacing the adjusted numbers of cases (with which the model was originally fit) with the original cumulative numbers of cases, and assuming only the Roche Elecsys assay (associated with the highest seroprevalence estimates) is used.

Uncertainty around our estimated proportions infected can come from both the model fit, and from the search for times to seroreversion and lead or lag between seroprevalence and reported cases. To characterize the uncertainty, we took the best (bottom) five percentile LOO median RMSEs across parameter combinations (times to seroreversion and lead or lag), estimated the proportion infected for the corresponding subset of models to include the 95% uncertainty intervals (UIs) around each model fit, and extracted the range of estimates for each point in time and state (including the 95% UIs). U.S.-wide uncertainty estimates do not include model uncertainty (which in any case was significantly smaller than that from selection of times to seroreversion and lead or lag), and were estimated by producing a mean seroprevalence and estimated proportion infected weighted by state populations, for each of the models in the best five percentile models, and then taking the range of values at each point in time.

Estimates of the proportions of state populations infected as per the best waning model by LOO median RMSE, and the corresponding uncertainty intervals, are provided, by state and week, in SI Dataset S2.

We combine the estimated proportion infected with vaccination coverage (what we refer to in the text as the “estimated proportion infected and/or vaccinated”, or EPIV) by making assumptions on the correlation between the two. We show results assuming an independent probability, such that the probability of being vaccinated has no bearing on the probability of having been infected (i.e., P(vacc) + P(inf) − P(vacc) × P(inf)). We also show results assuming a perfect negative correlation (P(vacc) + P(inf)).

To understand variation in the extent to which naive (not yet infected or vaccinated) individuals had been prioritized by vaccination campaigns, we defined the following metric,

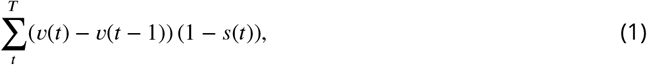

where *s*(*t*) is the EPIV (combined proportion infected and vaccination; see above) at time *t*, and *v*(*t*) is the vaccination coverage at time *t*. This metric estimates the proportion of the population that was vaccinated before being infected, assuming vaccinations were distributed independently of prior infection status.

We also repeated analyses with generalized additive models, with which we found statistically significant nonlinear relationships between covariates and seroprevalence (Figure S16). However, predicted seroprevalence values from the GAM were very similar to those predicted using the GLM (see Section “Accounting for non-linear relationships” in SI).

R v4.2 ***R Core Team*** (***2022***) was used in all analyses.

## Data Availability

All data used and produced will be available online.

## Acknowledgments

We would like to thank Harrell Chesson, Kristie Clarke, Jefferson Jones, Augustine Rajakumar, Heather Scobie, Jerome Tokars, and Ryan Wiegand for reviewing and helping improve the manuscript.

## Disclaimer

The findings and conclusions in this report are those of the authors and do not necessarily represent the official position of the Centers for Disease Control and Prevention. Use of trade names or commercial sources is for identification purposes only and does not imply endorsement by the U.S. Centers for Disease Control and Prevention.

