## Supplementary information for "Accounting for assay performance when estimating the temporal dynamics in SARS-CoV-2 seroprevalence in the U.S"

### Laboratory methods

Laboratory A used the Roche Elecsys Anti-SARS-CoV-2 pan-immunoglobulin immunoassay that targets the nucleocapsid protein and has a sensitivity of 100% (95% CI, 88.3%–100.0%) and specificity of 99.8% (95% CI, 99.7%–99.9%). Specimens were considered reactive at a cutoff index of 1.0 or greater without serum dilution (***US Food & Drug Administration, 2020b***; ***Bajema et al., 2021***). Laboratory B performed testing using the Abbott ARCHITECT SARS-CoV-2 IgG immunoassay targeting the nucleocapsid protein or Ortho-Clinical Diagnostics VITROS SARS-CoV-2 IgG immunoassay targeting the spike protein. Specimens tested by ARCHITECT were considered reactive at a cutoff index of 1.4 or greater, whereas specimens tested by VITROS were considered reactive at a cutoff index of 1.0 or greater. Using these definitions of reactivity, ARCHITECT had a sensitivity of 100.0% (95% CI, 95.8%–100.0%) and specificity 99.6% (95% CI, 99.0%–99.9%); VITROS had a sensitivity of 90.0% (95% CI, 76.9%–96.0%) and specificity of 100.0% (95% CI, 99.1%–100.0%; ***US Food & Drug Administration, 2020b***; ***Bajema et al., 2021***). For all assays, sensitivity was determined in symptomatic persons with real-time reverse transcriptase polymerase chain reaction–confirmed SARS-CoV-2 infection. The Roche assay was used through all survey rounds, the Abbott assay was used until July 2021 (round 24), and the Ortho assay was used until January 2021 (round 13). All assays were granted Emergency Use Authorization by the U.S. Food and Drug Administration and used according to the Instructions for Use provided by the manufacturers (***US Food & Drug Administration, 2020a***,b; ***Bajema et al., 2021***).

### Additional results

**Table S1.** Exponentiated regression coefficients (risk ratios) and standard errors of the binomial GLMs for the main reference model and best waning model (chosen by LOO RMSE; Table 1 and Fig 2 in the main text), in which the percentage of the population reported as a case ("Sqrt % cases") has been adjusted assuming 24, 42, and 97 weeks to seroreversion for Abbott, Ortho, and Roche assays, respectively, and a −1 week lag between seroreversion and a case being reported.

| | Main reference model | Best waning model |
|---|---|---|
| Sqrt % cases | 2.611*** (0.065) | 1.762*** (0.023) |
| Sqrt % deaths | 1.473** (0.269) | 11.569*** (1.685) |
| % excess deaths | 1.021 (0.261) | 3.442*** (0.834) |
| Sqrt % cases hospitalized | 0.967 (0.023) | 0.776*** (0.017) |
| Ln % tested (PCR) | 0.895*** (0.035) | 1.446*** (0.050) |
| % vaccinated | 0.998*** (0.0004) | 1.001*** (0.0004) |
| % Abbott Architect | 0.994*** (0.0002) | 0.997*** (0.0002) |
| % Roche Elecsys | 1.003*** (0.0002) | 1.002*** (0.0002) |
| % cases 0–19 years | 0.972*** (0.009) | 1.005 (0.009) |
| % cases 20–49 years | 1.022*** (0.007) | 1.017** (0.007) |
| % cases 50–69 years | 1.032** (0.015) | 1.070*** (0.016) |
| RMSE | 0.02171 | 0.01994 |
| LOO median RMSE | 0.02226 | 0.0204 |
| Observations | 1398 | 1398 |
| Log likelihood | −7482 | −7250 |
| Δ AIC | 464 | 0 |
| *Note:* | | *p<0.1; **p<0.05; ***p<0.01 |

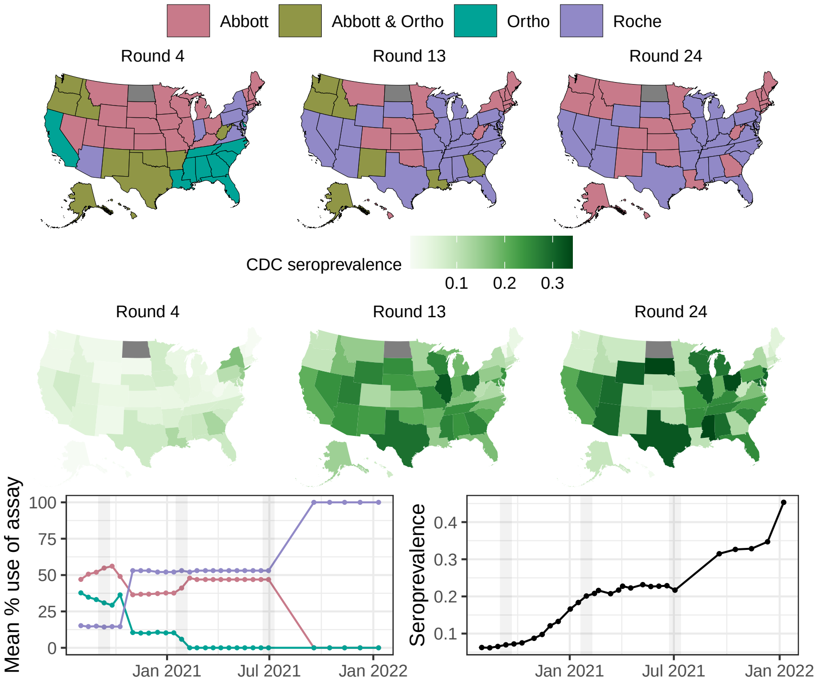


**Figure S1.** Spatial distribution of assays used in each state and seroprevalence, at three points in time, September 2020 (round 4), January 2021 (round 13), and July 2021 (round 24). Mean percentage use of each of the assays across states over time, and country-wide seroprevalence over time. The vertical gray bands in the time series indicate the three time points for the maps. Note that the distribution of assay use shown for July 2021 (round 24) remained the same between February and July 2021 (rounds 15 to round 24). After July 2021, all states used Roche assays exclusively (see ***Figure S3***).

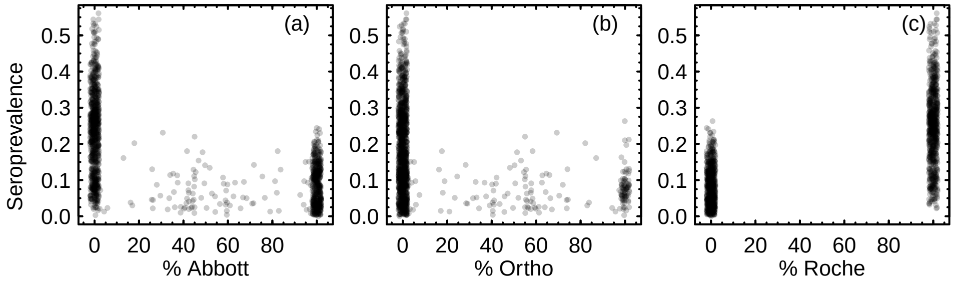


**Figure S2.** Seroprevalence from the CDC serosurveys as a function of the percentage of each assay used for each serosurvey, for (a) Abbott, (b) Ortho, and (c) Roche assays. Points that were equal to 0% or 100% on the x-axes have been jittered for greater clarity. Note that Ortho assays were only used until January 2021 (the first 13 rounds of the serosurveys).

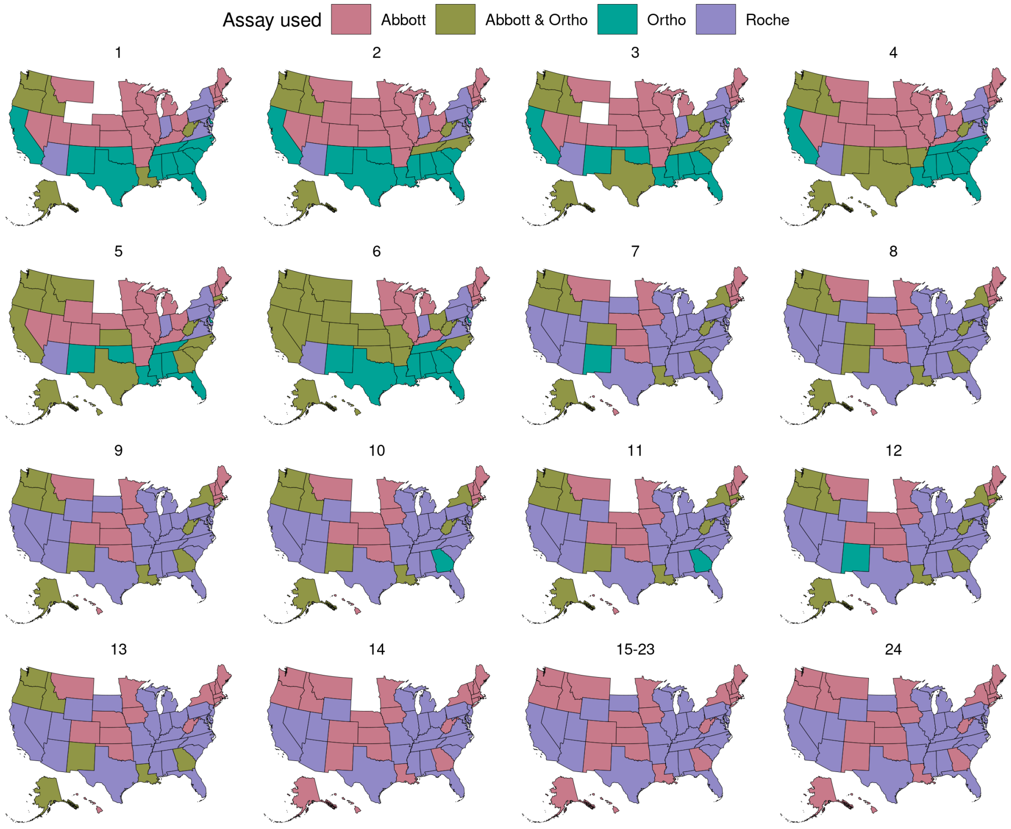


**Figure S3.** Assays used in each state, per round. After July 2021 (round 24), serosurveys in all states were performed only using Roche Elecsys assays.

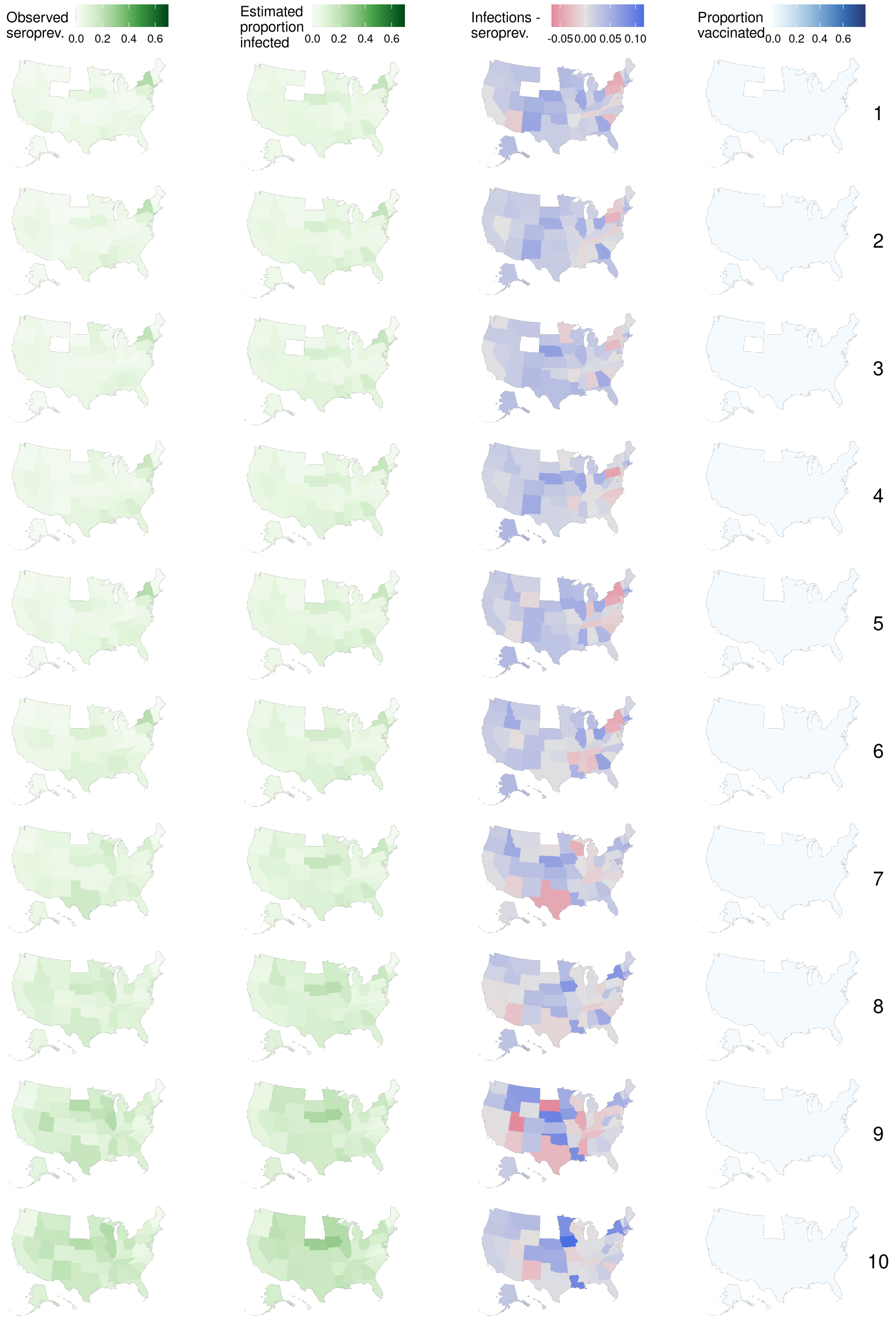


**Figure S4.** Observed seroprevalence, estimated proportion infected using the best waning model as per LOO median RMSE (Table 1 in the main text), the difference between estimated proportion infected and observed seroprevalence, and the proportion of the population vaccinated, between July 2020 and December 2020 (rounds 1–10 of the serosurveys).

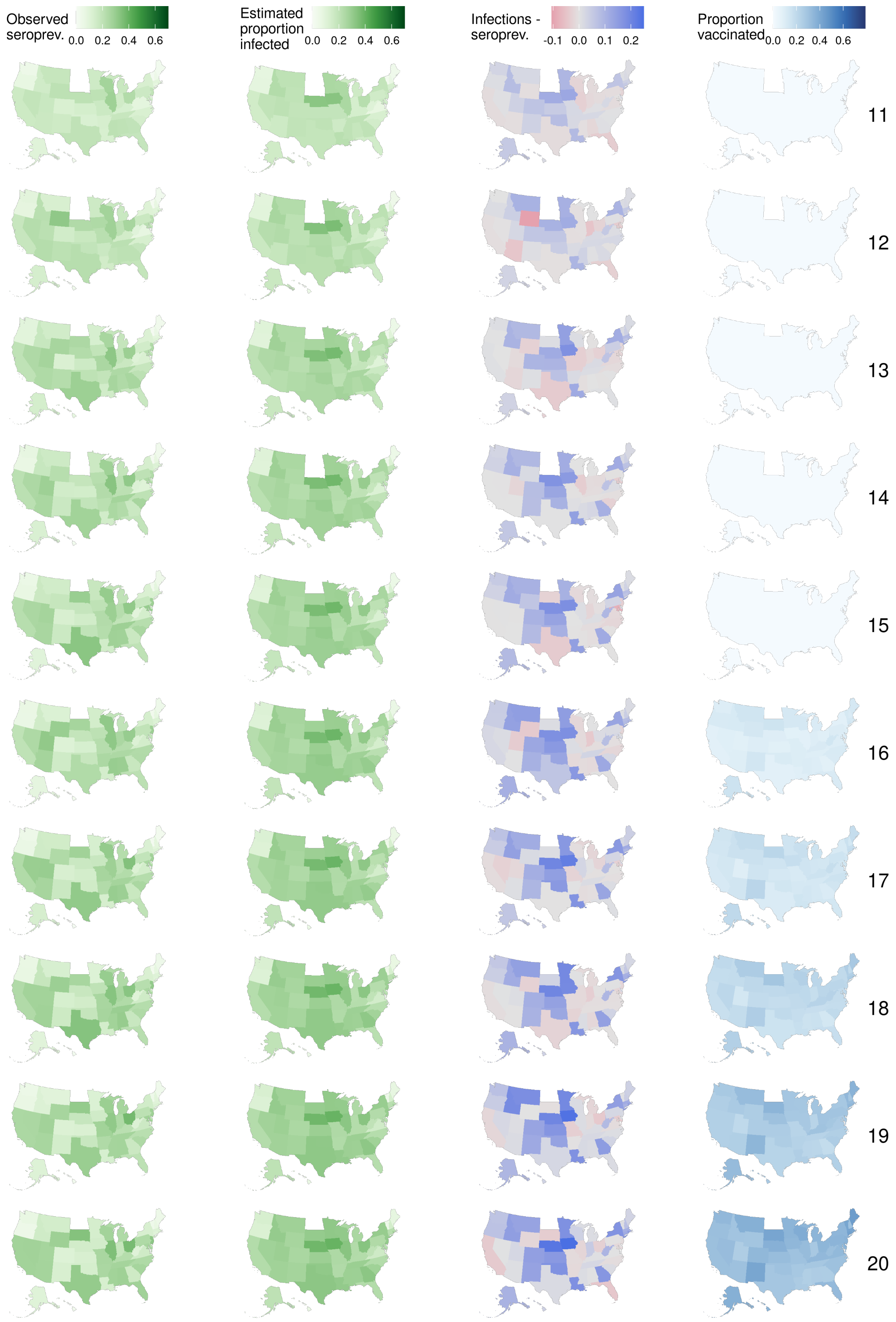


**Figure S5.** Observed seroprevalence, estimated proportion infected using the best waning model as per LOO median RMSE (Table 1 in the main text), the difference between estimated proportion infected and observed seroprevalence, and the proportion of the population vaccinated, between December 2020 and May 2021 (rounds 11–20 of the serosurveys).

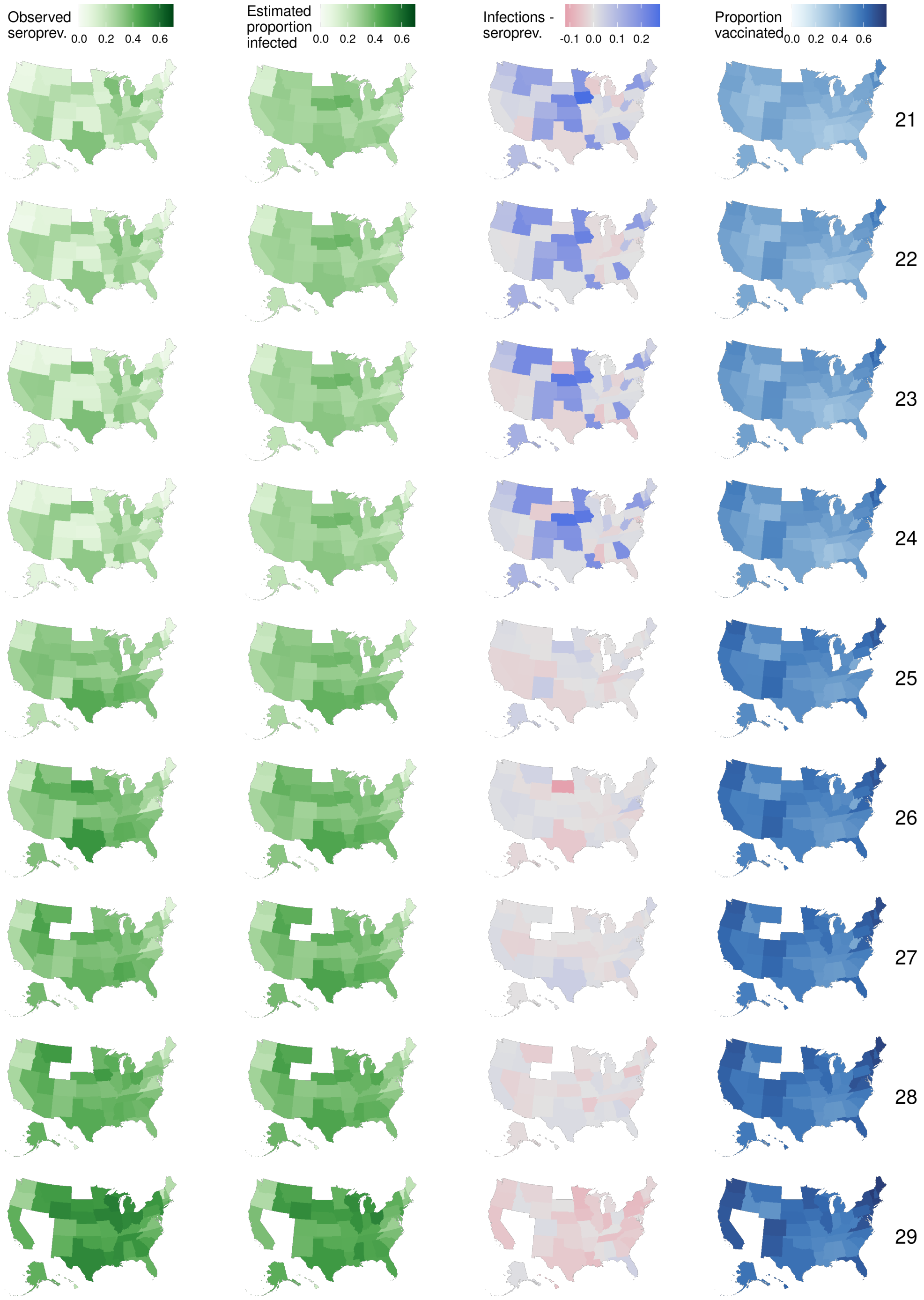


**Figure S6.** Observed seroprevalence, estimated proportion infected using the best waning model as per LOO median RMSE (Table 1 in the main text), the difference between estimated proportion infected and observed seroprevalence, and the proportion of the population vaccinated, between May 2021 and January 2022 (rounds 21–29 of the serosurveys).

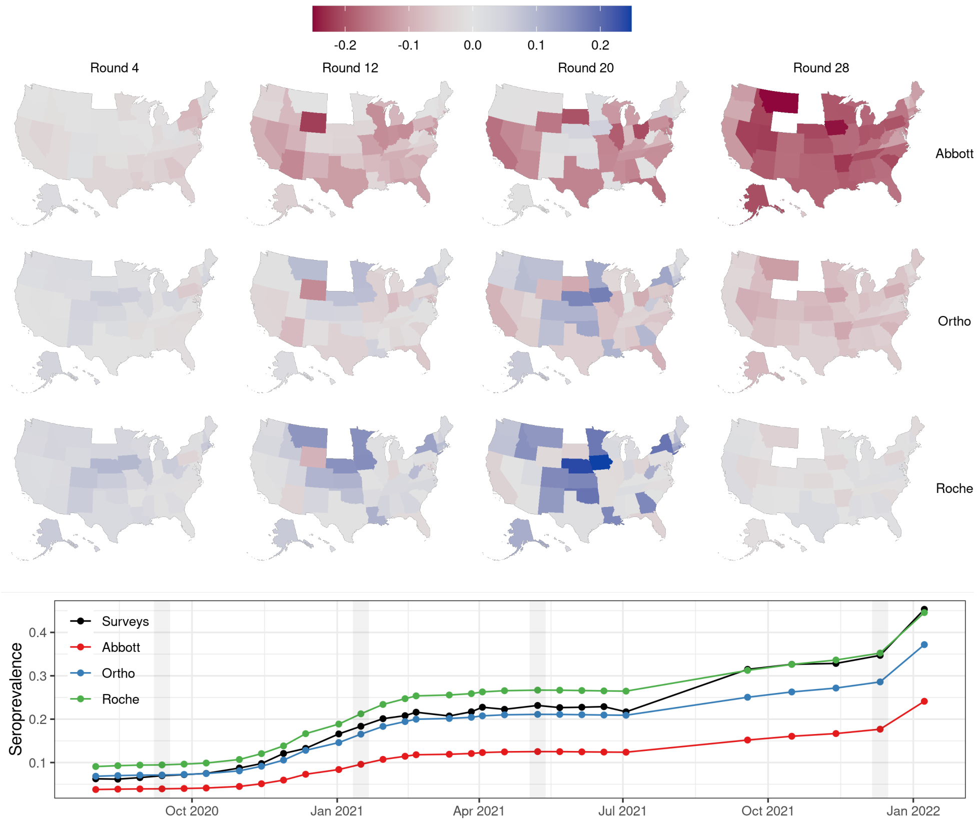


**Figure S7.** The differences in estimated seroprevalence were all states to have used the same assay, as estimated using the reference model. For the maps, assays are arranged in rows, and columns show four snapshots (survey rounds) in time. For example, in the top row, reds indicate the extent to which the seroprevalence would have been lower, had all states used Abbott assays only. Gray values mean that seroprevalence estimates would have changed little, because those states likely were already using those assays for those rounds. The time series in the bottom panel show the average seroprevalence across all states as observed in the surveys, and had all states exclusively used one of the assays. The four vertical gray lines approximately indicate the timing of the four snapshots in time of the maps above.

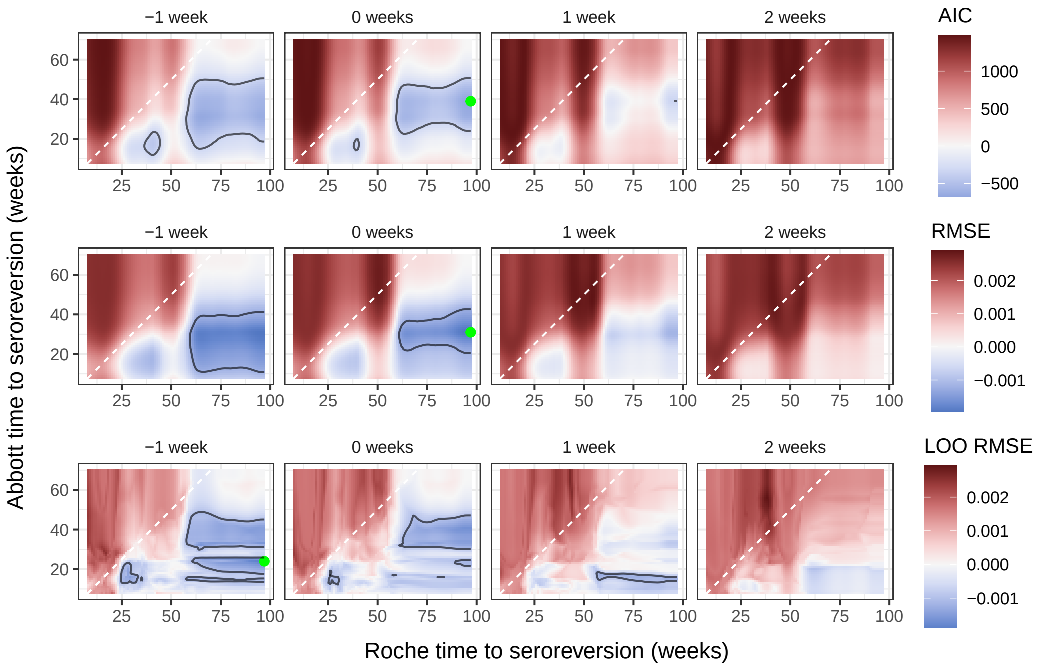


**Figure S8.** Model metrics for waning models compared to the (non-waning) reference model. Each row of panels is a different model metric (AIC, RMSE, leave-one-out median RMSE), and each column of panels refers to a different detection lead or lag. Within each tile plot, each pixel corresponds to a single model, where cases have been adjusted assuming three different times to seroreversion, one for each assay. Metrics are expressed relative to the metric for the main reference model (that does not account for waning); blues (respectively reds) indicate waning models that are better (respectively worse), per that metric, relative to the model without waning. Tile plots here show results for the subset for which the time to seroreversion for Ortho was at least 49 weeks (for AIC and RMSE) and 42 weeks (for LOO median RMSE). Green points indicate the best model by each metric, and contour lines enclose the best five percentile models as per each metric. Also see Table 1 in the main text.

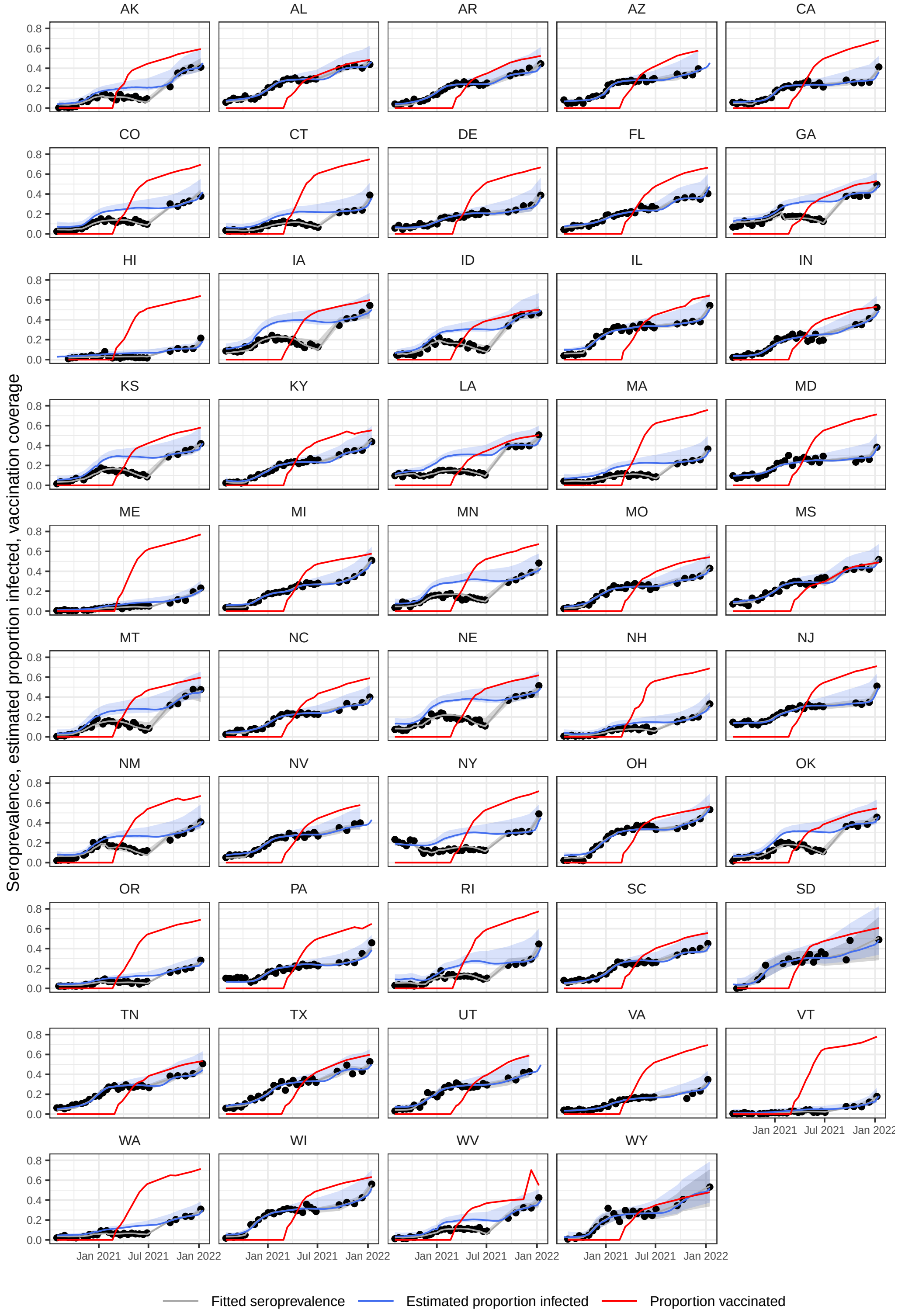


**Figure S9.** Serosurveys, fitted seroprevalence, estimated proportion infected from the best waning model (see Fig 2 and Table 1 in the main text), and vaccination coverage over time for each of the states included in the model. Confidence envelopes around fits and estimated proportions infected include model uncertainty and uncertainty around the selection of times to seroreversion and lead/lag between seroprevalence and reported cases.

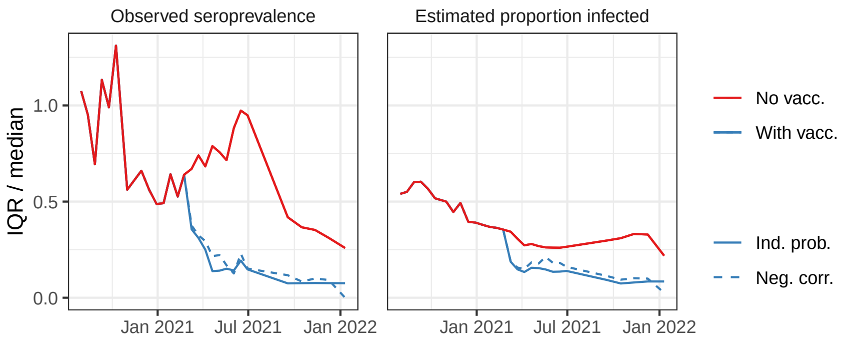


**Figure S10.** Spatial variation, expressed as the interquartile range (IQR) normalized by the median, in observed seroprevalence and estimated proportion infected, over time, across states. Solid blue lines assume the probabilities of being infected or vaccinated to be independent, while dashed lines assume a perfect negative correlation. The proportion infected were estimated using the best waning model (Table 1 in the main text). Smaller values of IQR / median point to greater homogeneity across the U.S. Accounting for vaccinations (blue lines), and the assays used and their different waning rates (right panel) both lead to greater homogeneity across states in seroprevalence and estimates of the proportion infected.

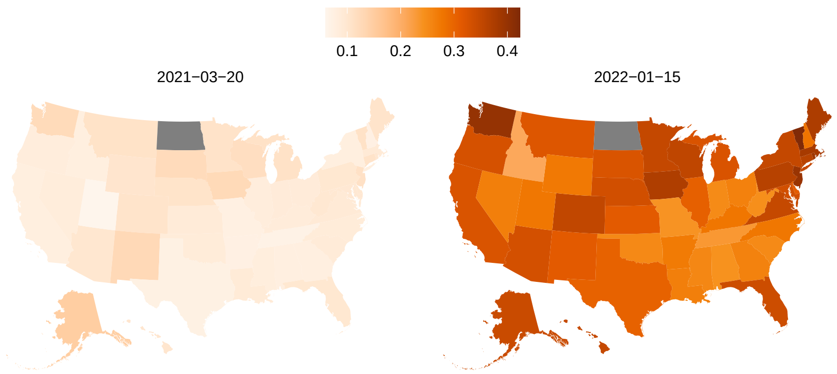


**Figure S11.** Pre-infection vaccination coverage at two different time points. The proportion of individuals that may have received a complete series of a vaccine prior to being infected, assuming that vaccinations were distributed independently of prior infection status (see Methods and Materials).

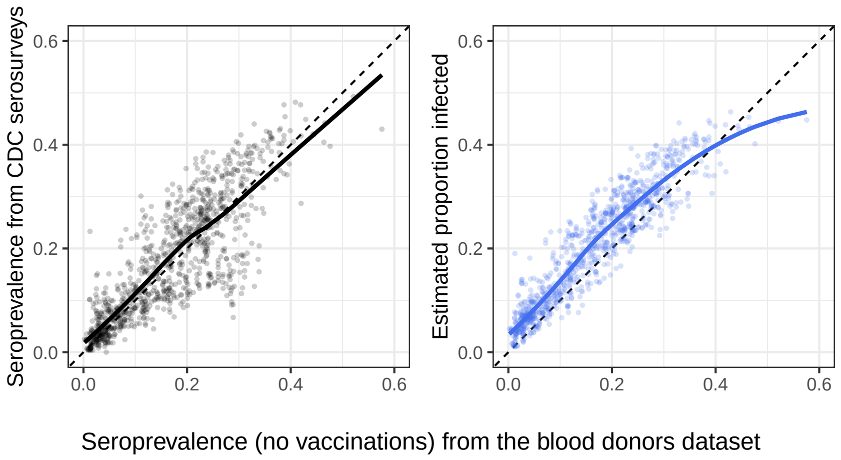


**Figure S12.** Comparison of estimated proportion infected (without vaccinations) from the blood donors dataset, and both the estimates from the serosurveys (black line and points) and estimated proportion infected (blue line and points) from the GLM model. Lines are LOESS fits to the underlying data.

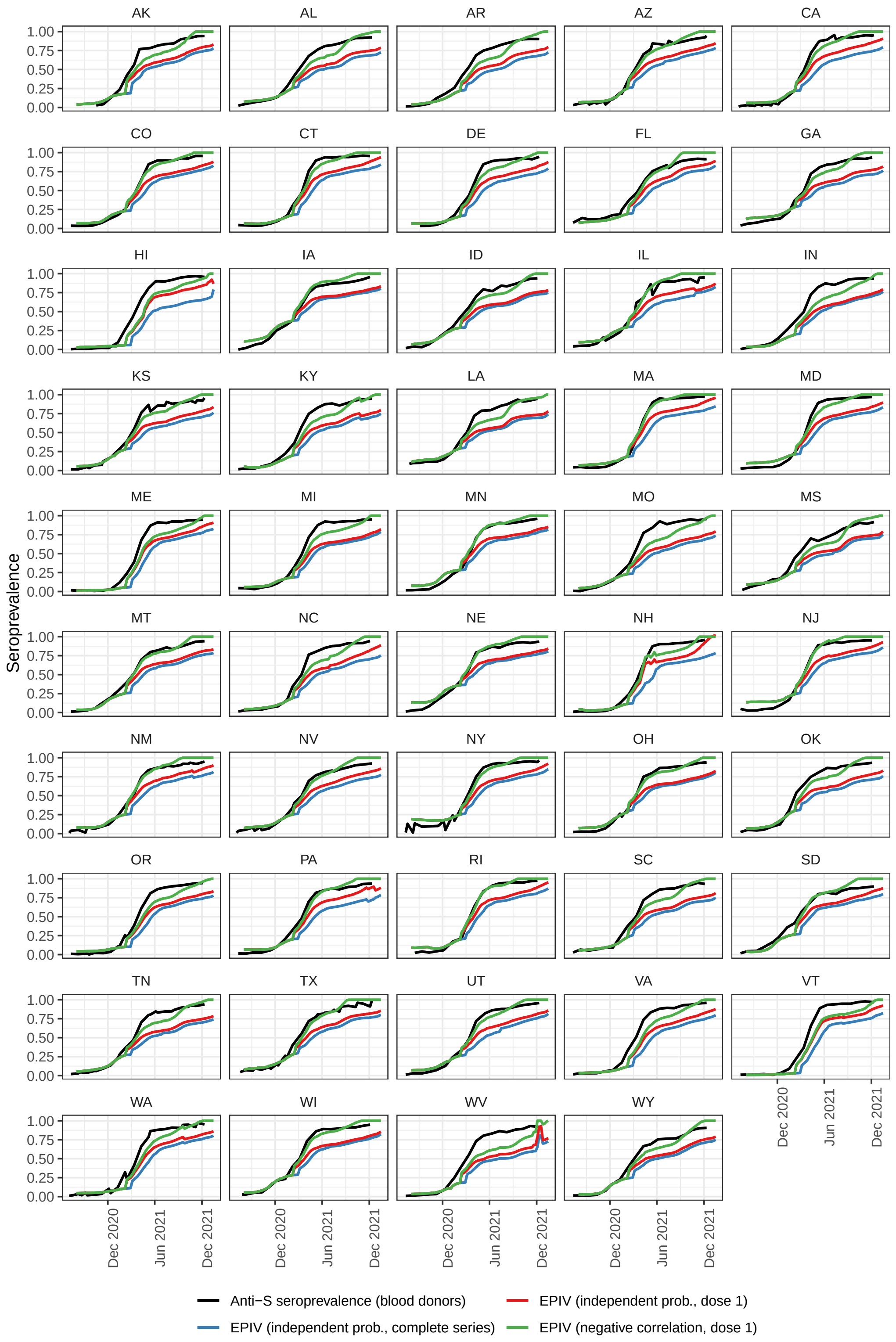


**Figure S13.** Time series of anti-S seroprevalence from the blood donors dataset, compared to estimated proportions infected and/or vaccinated using the best waning model (Table 1 in the main text).

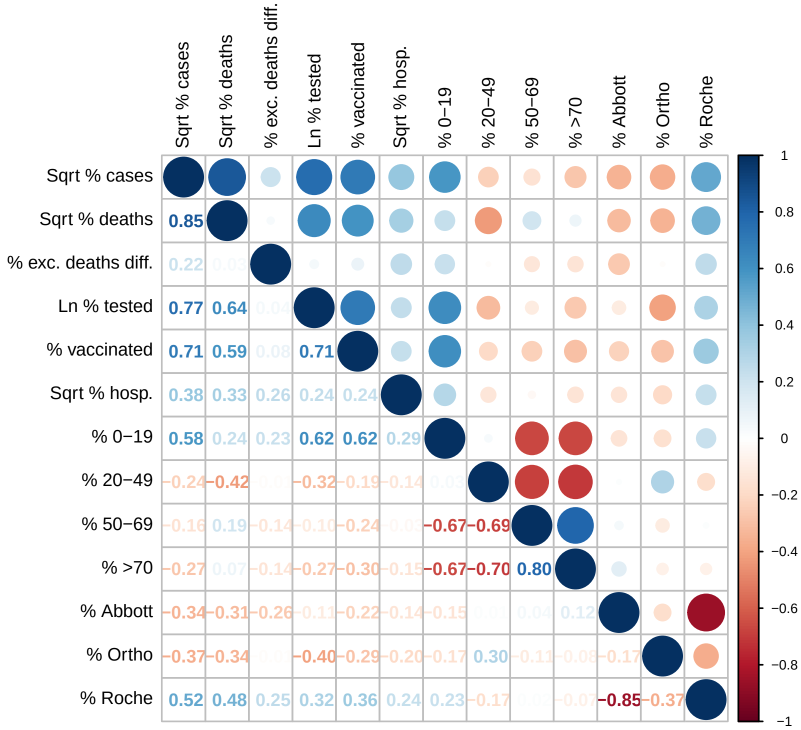


**Figure S14.** Pearson cross-correlations between variables used in the models. Note that "% >70" and "% Ortho" were excluded from the models. Darker blues and reds indicate larger positive and negative correlations between pairs of variables, respectively.

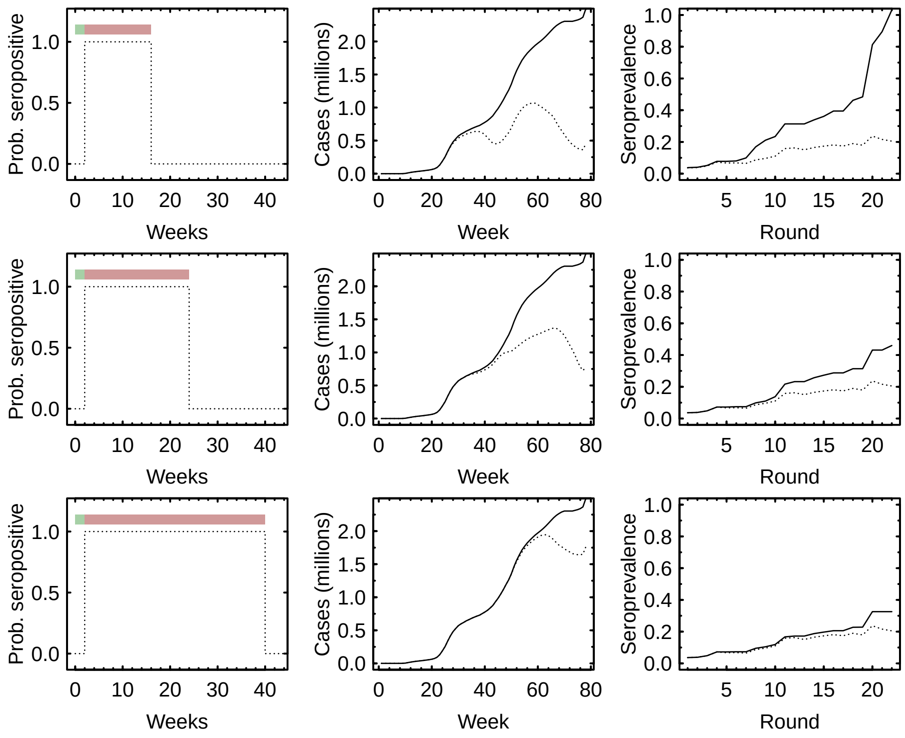


**Figure S15.** Rationale for adjusting cases. Each reported case is multiplied by a probability of testing seropositive (left column of panels), which takes the form of a step function, with a lead or lag in time between seroconversion and a case being reported (green bars), and a limited amount of time during which a case remains seropositive (red bars) before seroreverting. Using these step functions, cumulative cases can be adjusted to account for waning (middle panels). Here, solid lines are the cumulative cases, and the dashed lines are the cases adjusted for waning using the step functions in the left panels. The different step functions would imply different degrees to which serosurveys are an underestimate of the proportion infected (right panels). Here, solid lines are what the hypothetical proportion infected might have been, given the waning rates encoded in the step functions of the left column, and the dashed lines are the estimated seroprevalence (that include the effects of waning in antibodies). The top, middle, and bottom rows assume waning rates that go from faster to slower, so with faster waning rates (shorter times to seroreversion; top row), agreement between seroprevalence and the proportion infected is worse than if waning rates were slower (bottom row).

### Accounting for non-linear relationships

Acknowledging that the relationship between seroprevalence and some of the covariates may also be nonlinear, we ran the same analyses described above using generalized additive models (GAMs). In these models, all covariates except for the interaction between week and state, and the percentage of Roche tests, are smooth (where the dimensions of the bases are constrained to $k = 5$). Allowing the interaction between week and state to be smooth (and thus allowing the seroprevalence in each state to be described by a smooth function of time) did lead to models with better fits across all metrics. However, conferring such a degree of flexibility to the model would have meant that some of the signal due to waning that might otherwise have been captured through our adjusted cases would instead have been picked up in this interaction term. GAM models were fit using the 'gam' function in package 'mgcv' in R v4.2. All models were fit using REML, and we used a double-penalty shrinkage (`select = TRUE`).

Results show significant non-linearities in some of the relationships with covariates (***Figure S16***) but predicted seroprevalences from both GLM and GAM models were very highly correlated (***Figure S17***).

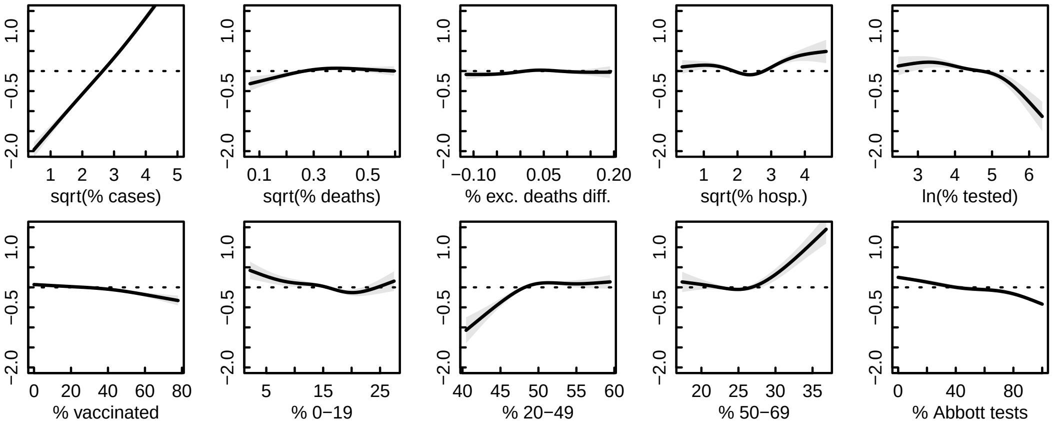


**Figure S16.** GAM functions for each of the predictor variables in the main reference model with a linear interaction between week and state. Note that the percent Roche tests used was included as a linear term because it only consists of either 0% or 100% values (see ***Figure S2***), and had a statistically significant positive slope.

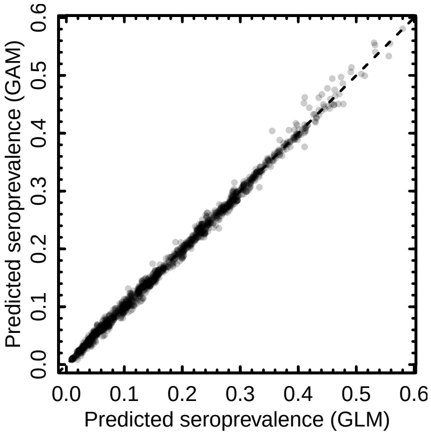


**Figure S17.** Comparison of predicted seroprevalences using the main reference model made by the GLM (used in the main text) and GAM model. The Pearson correlation coefficient is 0.998.

### Supporting Data

Data used to fit the GLM models (see Section "Data" in Methods and Materials) and estimates of proportions of state populations infected by week are provided as supplementary data files. Estimates are produced using the best waning model by leave-one-out median RMSE (see Table 1 and Figure 2 in the main text, and ***Figure S8*** above). See Section "Models" in Methods and Materials for how the lower and upper bounds we provide for these estimates are calculated.
